# GWAS Reveals Distinct Genetic Architecture of Schistosomiasis-Induced Hepatic Fibrosis with *DGKG* as a Key Mediator

**DOI:** 10.64898/2026.03.21.26348960

**Authors:** Miao Zhou, Chao Xue, Lichao Zhang, Yunyi Hu, An Ning, Lifu Wang, Jia Shen, Langui Song, Beibei Zhang, Jiahua Liu, Yao Liao, Zebin Chen, Jehangir Khan, Zhongdao Wu, Chunxiu Chen, Xi Sun, Xiaoying Wu, Miaoxin Li

## Abstract

**Background & Aims:** Schistosomiasis is a major cause of hepatic fibrosis in endemic regions, yet the host genetic determinants of disease progression remain poorly defined. We aimed to identify genetic drivers and underlying mechanisms of schistosomiasis-induced hepatic fibrosis.

**Methods:** We performed a genome-wide association study (GWAS) of 984 *Schistosoma japonicum*-infected individuals from hyperendemic areas in China followed by multi-omics integration and experimental validation to identify causal genes and fibrogenic pathways.

**Results:** Schistosomiasis-associated fibrosis exhibited a genetic architecture distinct from metabolic and viral liver fibrosis, supporting pathogen-specific mechanisms. Eight novel susceptibility loci were identified, including a genome-wide significant signal at 16p13 (rs73575170, *P* = 3.9 × 10⁻⁸). Integrative mapping linked these loci to 262 genes enriched in liver sinusoidal endothelial cells (*P* = 5.84 × 10⁻⁵) and sphingolipid metabolism pathways (*P* = 4.19 × 10⁻⁵). Notably, Diacylglycerol kinase gamma (*DGKG*, rs6762330, *P* = 4.37 × 10⁻⁶) emerged as a key candidate, with its expression in peri-granuloma and periportal hepatocytes strongly correlating with fibrosis severity (*r* = 0.816). *In vivo*, *Dgkg* knockout attenuated hepatic fibrosis and immunopathology while restoring cholesterol homeostasis, whereas *Dgkg* overexpression exacerbated fibrogenesis and increased TNF-α levels tenfold.

**Conclusions:** This study identifies *DGKG* as a key mediator linking lipid metabolism and immune signaling in schistosomiasis-induced fibrosis, uncovering a pathogen-specific genetic mechanism and providing a potential therapeutic target for infection-associated liver fibrosis.

**Impact and Implications:** This study defines a pathogen-specific genetic architecture underlying schistosomiasis-associated hepatic fibrosis, distinguishing it from metabolic and viral fibrotic liver diseases. By integrating GWAS analysis and *in vivo* functional validation, *DGKG* is identified as a key regulator linking lipid metabolism and immune-driven fibrogenesis. These findings highlight liver sinusoidal endothelial cells and sphingolipid signaling as critical cellular and molecular contexts for schistosomiasis-associated hepatic fibrosis. Beyond advancing mechanistic understanding of schistosomiasis, this work provides a genetic and functional framework for targeting *DGKG*-mediated pathways, with potential implications for precision therapies in parasitically mediated liver fibrotic disorders.

## INTRODUCTION

Schistosomiasis remains a significant global health challenge, with the World Health Organization (WHO) reporting that at least 251 million infected persons worldwide and approximately 700 million people are at risk in endemic regions (1, 2). Of particular clinical concern is the persistent progression of hepatic fibrosis in infected patients, which continues unabated even following timely administration of praziquantel--the current anthelmintic gold standard (3, 4). This insidious fibrotic process frequently culminates in cirrhosis and progression to advanced schistosomiasis, resulting in substantial impairment of both workforce productivity and quality of life (5). As of 2023, there are still 184,216 cases of schistosomiasis in China, including 27,768 cases of advanced schistosomiasis. However, there is currently a lack of effective drugs available for reversing schistosomiasis hepatic fibrosis, rendering a critical therapeutic gap in the prevention and treatment of schistosomiasis pathology. Therefore, it is imperative to elucidate the mechanisms underlying the occurrence and progression of schistosomiasis hepatic fibrosis, which will provide more precise scientific foundations for enhancing and optimizing strategies for the prevention and treatment of advanced schistosomiasis.

Notably, disease progression in schistosomiasis demonstrates significant interindividual heterogeneity, with only a subset of patients advancing to the advanced stage. Repeated infections, lifestyle behaviors, and other factors can accelerate the progression of schistosomiasis. However, population-based studies have shown differences in the pathological advancement of schistosomiasis at both group and individual levels. Of particular interest, African populations--despite millennia of schistosome exposure--exhibit attenuated hepatic fibrosis progression compared to non-African counterparts in endemic zones, suggesting potential evolutionary adaptations (6, 7). Furthermore, severe fibrotic manifestations display familial clustering in high-transmission areas, with highly infected households demonstrating spatial aggregation of advanced cases (8–10). This phenotypic variability also extends to Asian contexts. Schistosomiasis japonica has prevailed in the Yangtze River basin in China for over two millennia. Our field surveys in Jiangxi Province identified divergent clinical trajectories: rapid progression to advanced disease (characterized by severe peri-portal fibrosis, portal hypertension, and ascites) coexisting with non-progressors maintaining stable presentations. In addition to factors such as infection frequency, living environment, health status, and lifestyle, the host’s genetic factors may also play an important role in the progression and severity of schistosomiasis (11). In 1996, researchers first identified *SM1*, the quantitative trait locus regulating controls *Schistosoma mansoni* infection intensity (fecal egg count) through parametric linkage analysis (12). This region contains the typical Th2 cytokines such as IL-4, IL-5 (13), IL-9, IL-10, and IL-13 (14–16). Subsequent studies into advanced schistosomiasis susceptibility have mainly focused on the immunogenetic mechanisms of hepatic fibrosis or analyzed specific genetic regions through case-control experiments without a comprehensive exploration at the whole-genome level (17–20). Hence, existing data incompletely elucidate the genetic mechanisms influencing the progression of schistosomiasis.

To systematically investigate the genetic mechanism affecting hepatic fibrosis, the primary pathological damage of schistosomiasis, we collaborated with the Parasitic Disease Control Research Institute of Jiangxi Province, China, to conduct a three-year epidemiological survey in the counties surrounding Poyang Lake. We established strict sample inclusion criteria, recruited nearly a thousand schistosomiasis patients, collected blood samples and clinical imaging diagnostic data, and applied a GWAS (21) approach that does not rely on prior pathogenic knowledge. For the first time, we discovered multiple novel susceptibility loci significantly associated with fibrotic progression at genome-wide significance thresholds. Further, we verified the causal relationship between associated genes identified by post-GWAS and schistosomiasis hepatic fibrosis through *in vivo* experiments. This discovery of host genetic modifiers provides new insights into disease susceptibility, personalized medicine, and the development of new clinical drug targets.

## RESULTS

### Study population and clinical characteristics

We recruited 984 individuals with a history of *S. japonicum* infection from nine counties around Poyang Lake in Jiangxi province from 2015 to 2017, China, following the inclusion criteria outlined in the Methods section. Demographic information is presented in Table 1. The average age of the participants was 60 years, with a majority being male. Liver function tests showed high levels of hyaluronic acid (HA, 155.24 ± 181.07 ng/mL, mean ± SD; normal range < 120 ng/mL) and aspartate aminotransferase (AST, 34.32 ± 21.8 U/L, mean ± SD; normal range < 40 U/L) levels in the subjects (Table 1). The longer the exposure duration (Spearman’s *r* =0.46) and the higher the frequency of exposure to infested water (*r* =0.25), the more serious the liver damage and the higher the degree of hepatic fibrosis (Fig. 1a). In comparison, BMI (*r* = -0.19) and albumin (*r* = -0.32) exhibited a significant negative correlation with hepatic fibrosis (Bonferroni corrected *P* < 0.01) (Fig. 1a). Interestingly, approximately 3.25% of patients infected with *S. japonicum* did not develop hepatic fibrosis according to our data. Of the 289 patients exposed to infested water weekly (score 4), approximately 14% remained free of grade three hepatic fibrosis, ascites, and portal hypertension. However, among 366 patients with very low-frequency exposure (score 1), 2.5% progressed to advanced hepatic fibrosis, ascites, and portal hypertension (Supplementary Table 1). Therefore, genetic factors may contribute significantly to the progression of advanced schistosomiasis.

**Fig. 1.**
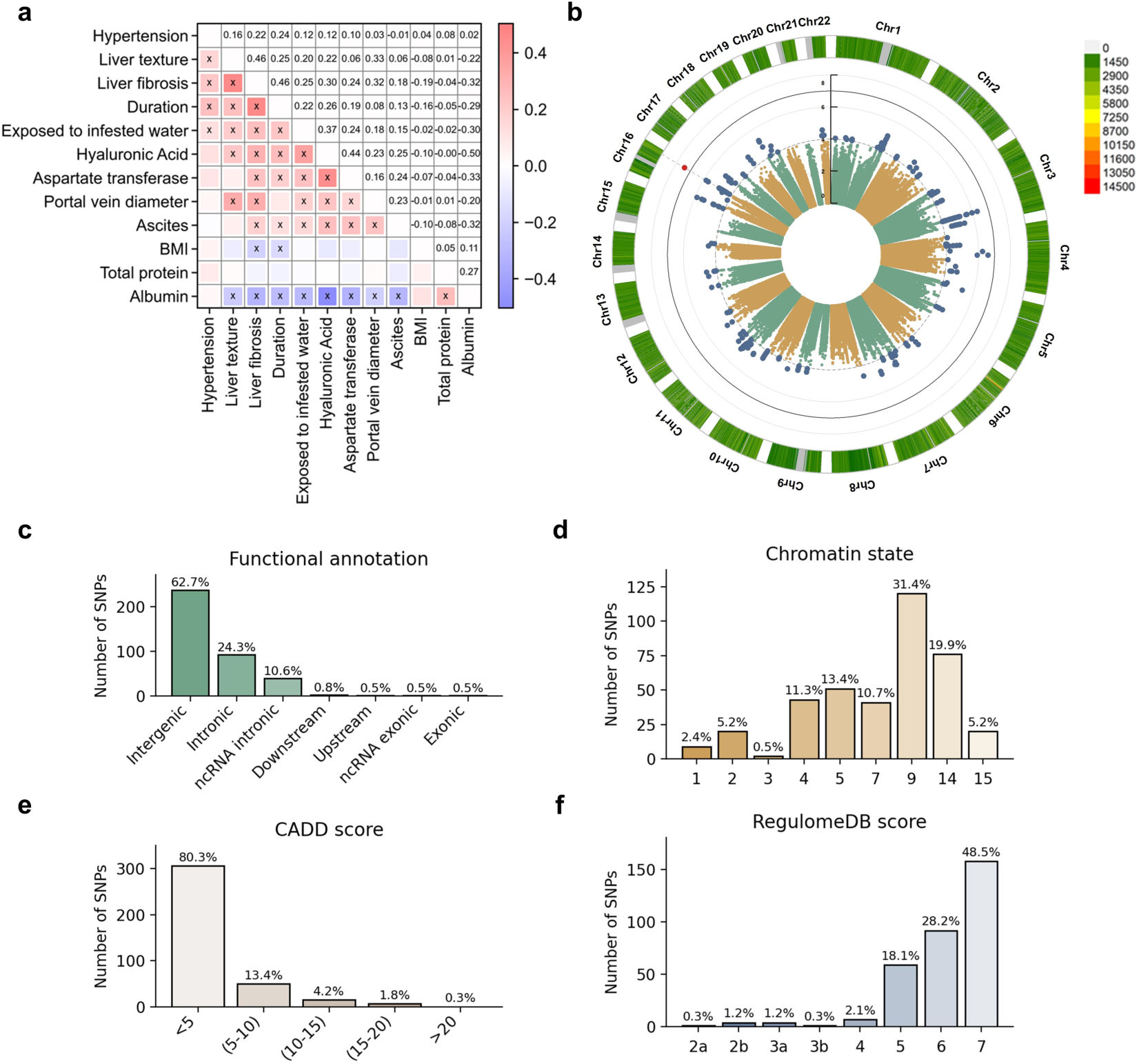
Correlation of clinical characteristics and SNP-based associations with schistosomiasis hepatic fibrosis in the GWAS analysis of n = 637 independent individuals. (a) Spearman correlation coefficient heat map among clinical characteristics. The color indicates the size of the correlation coefficient, and the “x” sign indicates that it reaches the significance level after Bonferroni multiple testing correction, that is, *P*<0.01/((12-1)*12/2), which is approximately equal to 1.5e-4. (b) Manhattan plot for genome-wide association analysis of schistosomiasis hepatic fibrosis. The y-axis shows the negative log10-transformed SNP two-sided *P* value from a linear additive model, and the x-axis is the base-pair position of the SNPs on each chromosome. The solid black line circle indicates a genome-wide significant *P* value threshold (*P*<1e-7) derived from Bonferroni correction, and the dashed gray line circle indicates a suggestive threshold (*P*<1e-5). (c), Distribution of SNP functional sequences in genomic risk loci from GWAS analysis results. (d, e, and f), Distribution of RegulomeDB categories, CADD scores, and min- chromatin state of all annotated SNPs in linkage disequilibrium (*r^2^* ≥ 0.6) with one of the GWS SNPs, respectively.

**Table 1.** Clinical characteristics of 984 schistosomiasis patients in this study.

| Characteristics | Values |
| --- | --- |
| Age, mean ± SD | 61.13 ± 11.48 |
| Female Sex — n. (%) | 38.70% |
| Smoking — n (“NO”, %) | 598 (60.8) |
| Alcohol drinking — n (“NO”, %) | 770 (78.3) |
| Duration, mean ± SD | 34.98 ± 16.37 |
| Score of exposure to infested water — n (%) |  |
| 1 | 366 (37.2) |
| 2 | 160 (16.26) |
| 3 | 169 (17.17) |
| 4 | 289 (29.37) |
| BMI median kg/m<sup>2</sup> (IQR) | 22.20 (20.53-24.35) |
| Hyaluronic Acid ng/mL, mean ± SD | 155.24 ± 181.07 |
| Portal vein diameter n (“Normal”, %) | 745 (75.71) |
| AST U/L, mean ± SD | 34.32 ± 21.80 |
| ALB g/L, mean ± SD | 47.38 ± 5.51 |
| Total protein g/L, mean ± SD | 79.49 ± 7.38 |
| Central obesity n (“NO”, %) | 619 (62.91) |
| Hypertension n (“NO”, %) | 523 (58.1) |
| Hepatic Fibrosis Grade n (%) |  |
| 0 | 32 (3.25) |
| 1 | 103 (10.47) |
| 2 | 128 (13.01) |
| 3 | 721 (73.27) |
| Ascites Grade n (%) |  |
| 0 | 890 (90.45) |
| 1 | 65 (6.61) |
| 2 | 20 (2.03) |
| 3 | 9 (0.91) |

### Genome-wide association analyses

We genotyped 900 individuals using the Illumina Asian Screening Array and identified 637 unrelated individuals based on the coefficient of population kinship (Methods section). After stringent quality filtering and genotype imputation, we retained 4,523,335 SNPs in the cohort of 637 unrelated individuals. Subsequently, we performed genome-wide association analysis using a linear regression model on this cohort (Fig. 1b), adjusting for the top seven ancestry principal components (PCs) and seven environmental factors (age, sex, etc. in Table 1) as covariates. We captured locus 16p13 reaching the genome-wide significance threshold (*P* = 1.65 × 10^-7^, according to the effective number of independent SNPs (22), see details in the methods section) in the cohort of 637 independent individuals. The Q-Q plot suggested no overall inflation of the genome-wide statistical results (λ = 1.008, Supplementary Fig. 1).

### Significant independent loci associated with schistosomiasis hepatic fibrosis

We identified eight genomic risk loci (*P* < 1 × 10^−5^, Supplementary Table 2), including nine significant independent SNPs (Table 2) and eight lead SNPs. We performed functional annotation of all candidate SNPs in the eight associated risk loci, revealing that 62.7% were intergenic and 24.3% were intronic. Additionally, 45% of these SNPs were located in regions enriched for chromatin states 1-7, suggesting potential regulatory roles in active transcription (Fig. 1c, d and Supplementary Table 3). A high probability of deleterious variant effect (Combined Annotation Dependent Depletion (CADD) score ≥15) was identified at eight loci (Fig. 1e and Supplementary Table 3), including rs117413297 (*SDAD1*), rs35340520 (*PPL*) and five SNPs in *WWOX*. The SNP rs117413297 (standard *beta* for the C allele = -0.287; 95% CI: -0.39 to -0.18; *P* = 5.52 × 10^-7^; Fig. 2a) in the exonic region of *SDAD1* is a synonymous variant (Table 2). RegulomeDB scores were less than six for 23.2% SNP, suggesting a high probability that these SNPs have a regulatory function (Fig. 1f).

**Fig. 2.**
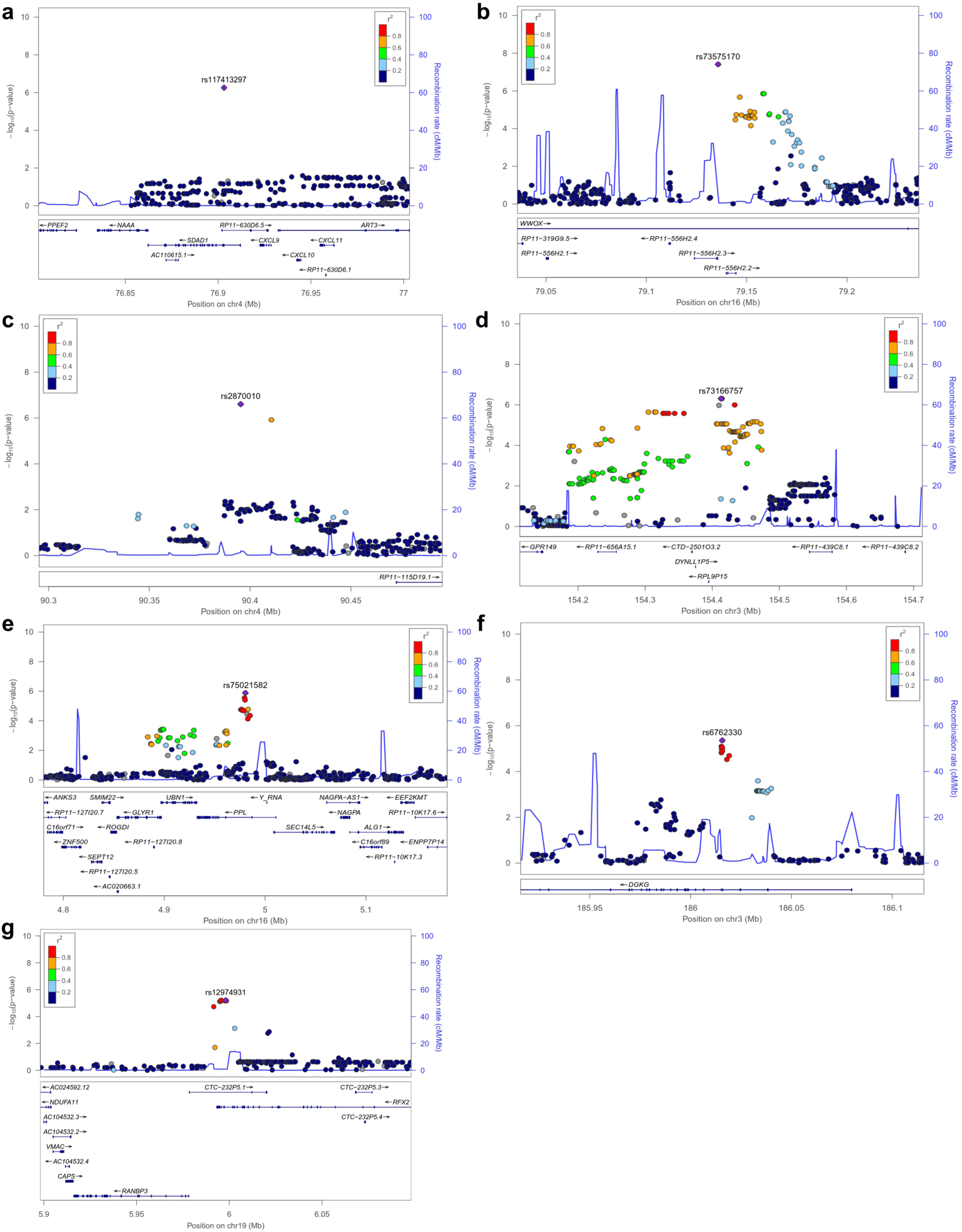
Regional plots for novel association loci. The x-axis represents the chromosomal position, and the y-axis is the –log10 of the *P* value of association. The index SNPs are shown as purple diamonds. Colors indicate the LD level with the index SNP: red (*r^2^* ≥ 0.8), orange (0.6 ≤ *r^2^* < 0.8), green (0.4 ≤ *r^2^* < 0.6), light blue (0.2 ≤ *r^2^* < 0.4), and dark blue (*r^2^* < 0.2). Blue lines represent recombination hotspots estimated from the East Asian population (from the 1000 Genomes Project, November 2014). Genomic positions are based on human genome assembly hg19. The plot shows the names and locations of the genes; the transcribed strand is indicated by an arrow. The Index SNPs include (a) rs117413297 at chromosome 4q21.1; (b) rs73575170 at chromosome 16q23.1; (c) rs2870010 at chromosome 4q22.1; (d) rs73166762 at chromosome 3q25.2; (e) rs75021582 at chromosome 16p13.3; (f) rs6762330 at chromosome 3q27.3; (g) rs12974931 at chromosome 19p13.

**Table 2.**
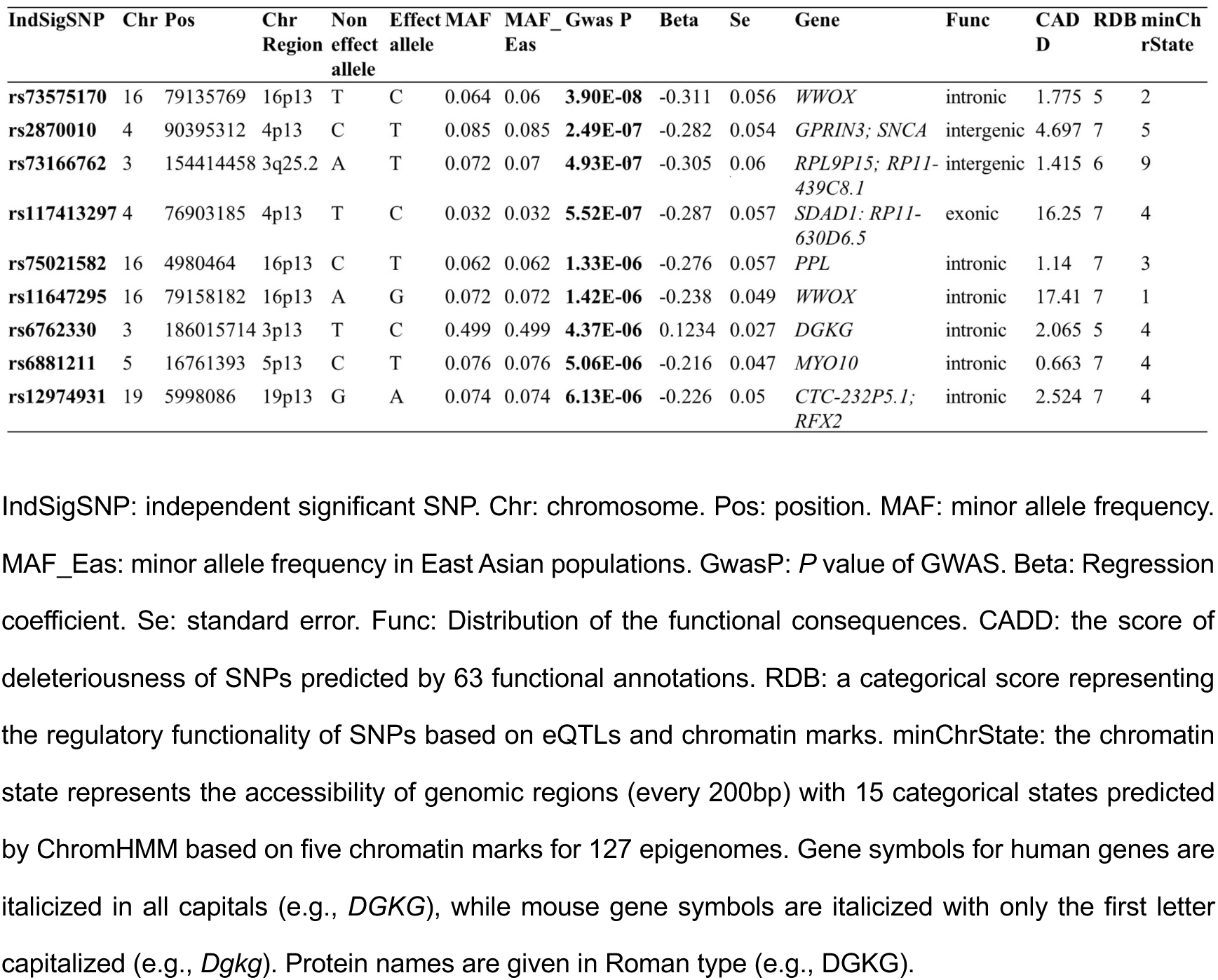
Association results for the independent significant SNPs in schistosomiasis hepatic fibrosis.

Notably, we found a genome-wide significant independent locus (*P* < 1.65 × 10^-7^) associated with hepatic fibrosis at chromosome 16p13 (lead SNP rs73575170; standard *beta* for the C allele = -0.311; *P* = 3.9 × 10^−8^; Fig. 2b and Table 2), located within an intron of the *WWOX* gene. The lead SNPs rs2870010 (P = 2.49 × 10^−7^) at 4p13 and rs73166762 (P = 4.93 × 10^−7^) at 3p25.2 in the risk loci are located in intergenic regions (Fig. 2c, d and Table 2). Loci detected in the intronic regions of the *PPL* (lead SNP rs75021582, standard *beta* for the T allele = -0.2762; *P* = 1.33 × 10^-6^; Fig. 2e) and *DGKG* (lead SNP rs6762330, standard *beta* for the C allele = 0.1234; *P* = 4.37 × 10^-6^; Fig. 2f and Table 2) genes also exhibited suggestive associations. As an eQTL, the T allele of rs6762330 is associated with the gene expression of the *DGKG* in blood tissue (*beta* = -0.12304, *P* = 0.0078; Supplementary Table 4). Additionally, as an mQTL, the C allele of rs6762330 affects the methylation level of the *DGKG* gene region (*beta* = - 0.016, *P* = 1.91 × 10^−11^; Supplementary Table 4) (23). Loci in 19p13 (lead SNP rs12974931, standard *beta* for the A allele = 0.074; *P* = 6.13 × 10^-6^; Fig. 2g and Table 2) in the intron of the *RFX2* gene. A detailed catalog of variants in the associated genomic loci is provided in Supplementary Table 3. We found that the regression coefficients (*beta*) of significant SNPs in genomic risk loci were predominantly negative (180/190, Supplementary Table. 3), indicating a milder hepatic fibrosis phenotype in individuals carrying the minor alleles (frequency < 10%).

To investigate whether the genetic loci associated with schistosomiasis-induced liver fibrosis are also implicated in liver fibrosis of other etiologies, we systematically retrieved 13 published GWAS association data on hepatic fibrosis from the GWAS Catalog (24–35) and compared the associated loci with those identified in our study (*P* < 1×10^−5^). Strikingly, we observed no overlapping associated loci or genes between our GWAS results and those associated with other liver fibrosis etiologies, such as NAFLD, viral hepatitis, and alcohol-related disease (Supplementary Table 5). This lack of overlap underscores the distinct genetic architecture of schistosomiasis hepatic fibrosis, further emphasizing the novelty and disease-specific nature of our findings.

### Implicated genes in schistosomiasis hepatic fibrosis

To link the variants significantly associated (*P* < 1×10^−5^) with schistosomiasis liver fibrosis to genes, we applied three gene-mapping strategies implemented in Functional Mapping and Annotation (FUMA) (36): positional mapping, eQTL mapping, and chromatin interaction (CI) mapping (Methods). Positional gene mapping aligned SNPs to 17 genes by genomic location, eQTL gene mapping linked cis-eQTL SNPs to 29 genes whose expression they influence, and chromatin interaction mapping annotated SNPs to 254 genes based on 3D DNA-DNA interactions (Supplementary Table 6). Among these, 38 genes were identified by associated with at least two mapping strategies, and five were supported by all three SNP-gene matching strategies (i.e., *WWOX*, *DGKG*, *PPL*, *UBN1*, *GLYR1*, (Fig. 3a)).

**Fig. 3.**
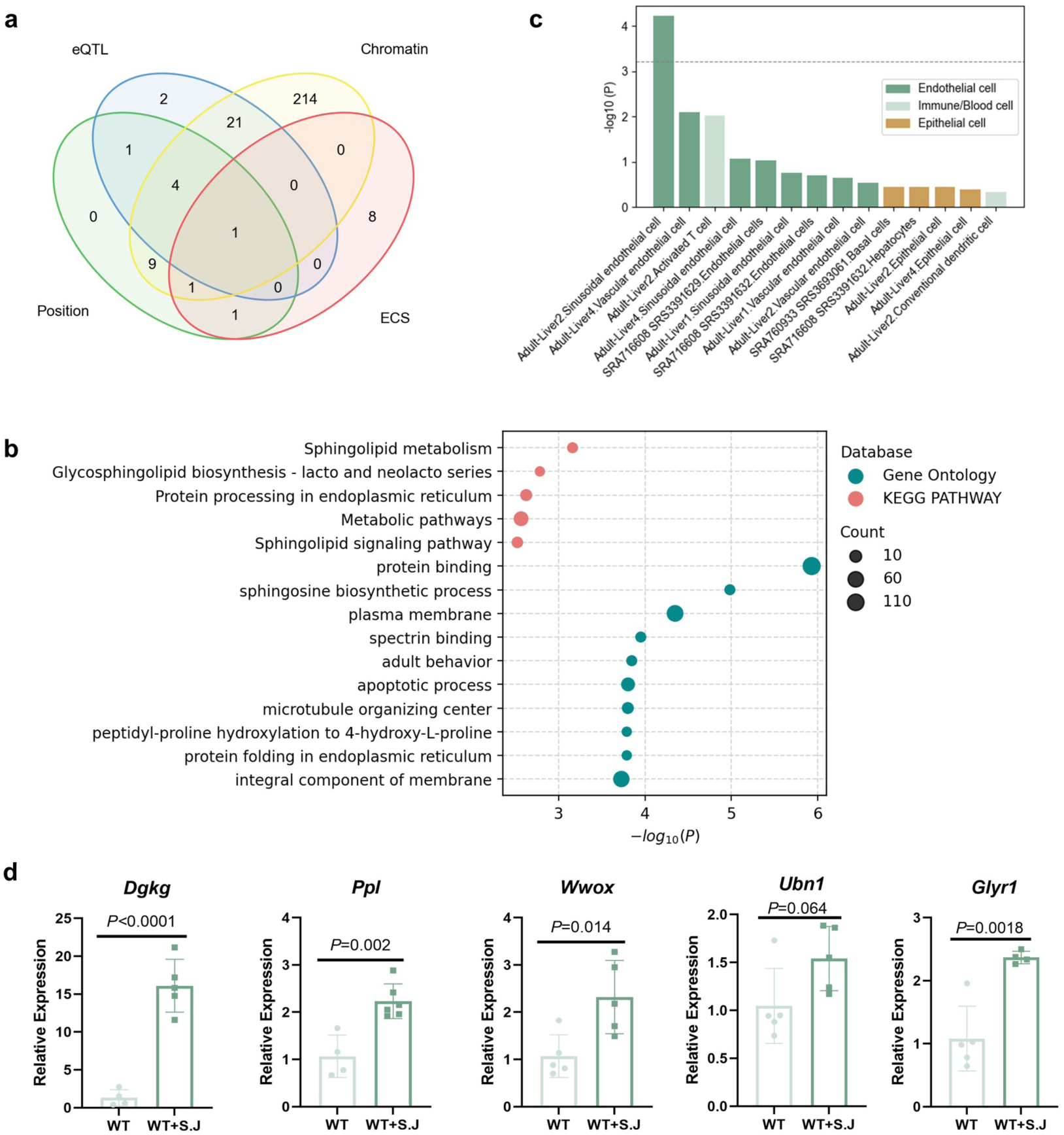
Implicated genes, pathways, and cell types for schistosomiasis hepatic fibrosis. (a) Venn diagram showing overlap of genes implicated by positional mapping, eQTL mapping, chromatin interaction mapping, and ECS. (b) GO and KEGG pathway enrichment for associated genes (ECS *P*<0.01). (c) Associated cell types in the liver. A total of 82 single cell types were tested in the liver, but due to image size limitations, only the top 15 are displayed. The dashed line represents the Bonferroni corrected significance level of 6 × 10^-4^. (d) Expression of candidate genes in the liver of mice infected with *S. japonicum* for 8 weeks (8 samples in each group). Bars represent the mean and error bars represent the mean ± SD. Statistical analysis was performed using a two-sided Student’s T-test.

We focused on the *WWOX* gene and found that some of its loci have been reported to be significantly associated with falciparum malaria (the most significant *P* = 3.72 × 10^−21^, UKBB, http://www.nealelab.is/uk-biobank/), eosinophil counts (37) (the most significant *P* = 7 × 10^-11^), pulmonary fibrosis (the most significant *P* = 6.39 × 10^−21^, UKBB), alcoholic hepatitis (the most significant *P* = 3.34 × 10^−21^), and cholangitis (the most significant *P* = 3.7 × 10^−19^, UKBB) in PhenoScaner (38) (Supplementary Table 7). Gene *PPL*, *UBN1*, and *GLYR1* are located at the same risk locus on chromosome 16p13.3 and are associated with BMI and cancer. Loci within the *PPL* gene have been reported to be associated with body height (the most significant *P* = 3×10^-86^), and liver enzyme levels (the most significant *P* = 3 × 10^−9^) (39, 40). Phenotypes associated with variants in *DGKG*, as reported in the GWAS catalog, are primarily related to anthropometric indicators such as BMI (the most significant *P =* 1 × 10^-50^), open-angle glaucoma (the most significant *P =* 8×10^-56^) and high-density lipoprotein cholesterol (HDL-CH, the most significant *P =* 2 × 10^-46^) levels (41–43).

Genome-wide association analysis identified multiple loci associated with schistosomiasis hepatic fibrosis. However, linkage disequilibrium (LD) may cause non-susceptible loci to show significant *P*-values alongside susceptible loci, potentially resulting in misleading results. To address this, we performed a genome-wide gene-based association study (GWGAS) analysis using ECS (44) to identify independent disease-associated genes. The results showed that *RFX2* (*P* = 2× 10^-5^), *SDAD1* (*P* = 5.8× 10^-5^) and *DGKG* (*P* = 2.2 × 10^-4^), which are also mapped in FUMA, were independently associated with hepatic fibrosis (Supplementary Table 8). Therefore, the *DGKG* was the only gene that was co-captured by ECS and three mapping strategies of FUMA (Fig. 3a).

We utilized the KOBAS database to perform GO enrichment and KEGG pathway enrichment for genes with an ECS *P*-value less than 0.01. Our findings revealed that the sphingolipid metabolism-related process was the most significantly enriched biological process (*P* = 4.19×10^-5^, Fig. 3b). This is consistent with previous research suggesting a close association between sphingosine and fibrosis in various tissues (45). The enriched pathways provide further strong evidence supporting the GWAS findings.

### Implicated driver cell types of liver fibrosis by the integrative analyses of GWAS and gene expression

Previous studies have shown that pathogenic genes tend to have higher selective expression in the pathogenic tissue for complex diseases (46, 47). We utilized GWAS summary data to estimate the relevant cell types involved in hepatic fibrosis using the PCGA (22, 44) platform. Our results revealed that the liver sinusoidal endothelial cell (LSEC) is the most relevant cell type for hepatic fibrosis (*P* = 5.84 × 10^-5^, Fig. 3c, Supplementary Table 9), surpassing the Bonferroni-corrected significance threshold of 6 × 10^-4^ (0.05/82, Methods section). This finding is consistent with the known pathogenesis of hepatic fibrosis in schistosomiasis. The fenestrae of LSECs disappear and form an organized basement membrane, which is the prelude to fibrosis (48). The LSECs primarily contribute to the hepatic fibrosis process by mediating liver inflammatory response, participating in sinusoidal angiogenesis and vasculogenesis, activating liver stellate cells, secreting various pro-inflammatory cytokines, participating in extracellular matrix generation, and mediating liver microcirculatory disturbance (49).

### Increased expression levels of candidate genes in schistosomiasis hepatic fibrosis

To validate the genes discovered by GWAS, we measured the expression levels of candidate genes in the livers of mice infected with *S. japonicum*. We analyzed the expression of three candidate genes (*Dgkg*, *Ppl*, and *Wwox*) in the total liver RNA of mice infected with *S. japonicum* in the eighth week. Our results showed significant up-regulation of these three candidate genes in the infected liver tissue (*P* < 0.05/3, Fig. 3d), suggesting their potential involvement in schistosomiasis development. In particular, the average expression of the *Dgkg* gene in the infected mouse livers was 16-fold that of the uninfected group (*P* < 0.0001, Fig. 3d).

### DGKG is expressed in hepatocytes near the periphery of granuloma and blood vessels

To investigate the expression of *Dgkg* in the development of schistosomiasis, we detected the mRNA levels of *Dgkg* in the liver tissues of mice infected with *S. japonicum* at different time points (4, 6, 8, 10, and 14 weeks). The expression of *Dgkg* is dynamically regulated following *S. japonicum* oviposition (3-4 weeks), peaking at week 8 post-infection and subsequently downregulating over time (Supplementary Fig. 2). Pearson correlation analysis, revealed a strong correlation between the expression of *Dgkg* and the fibrosis marker gene *Acta2* in the liver tissues of infected mice at different stages of infection (Fig. 4a, *r* = 0.816, *P* < 0.0001).

**Fig. 4.**
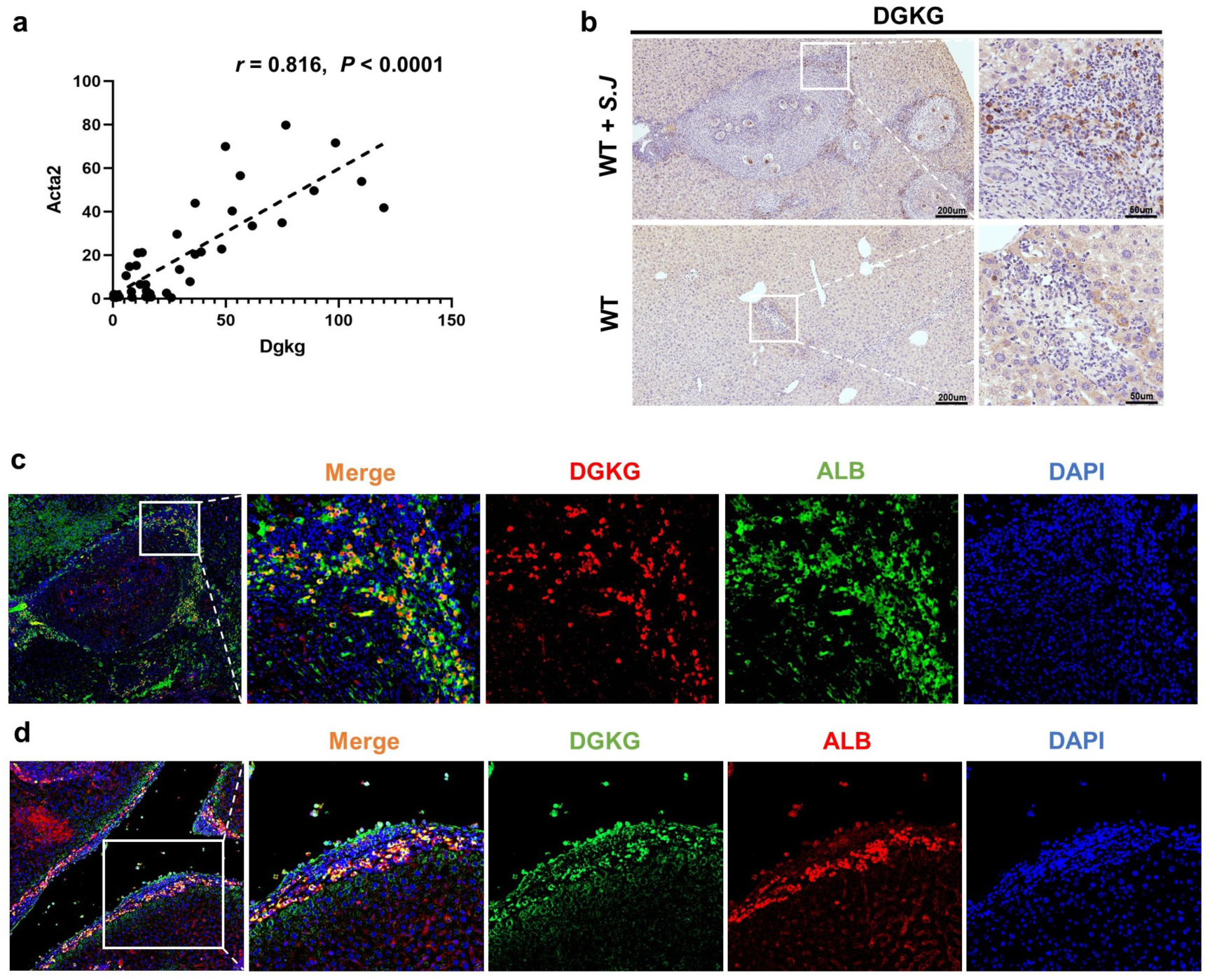
Expression of DGKG in schistosomiasis hepatic fibrosis. (a) Pearson correlation between *Dgkg* and *Acta2* expression in mice liver tissues at different infection time points (4, 6, 8, 10, 14 weeks, 7-8 samples in each infection time point). (b) Immunohistochemical staining results of DGKG in liver tissues of *S. japonicum* infected and normal mice. (c) Immunofluorescence co-staining results of DGKG co-localized with Albumin surrounding the granulomas. (d) Immunofluorescence co-staining results of DGKG co-localized with Albumin in perivascular hepatocytes. The IHC and IF images show the liver tissue of mice infected with *S. japonicum* for 8 weeks.

We then examined the expression region of *Dgkg* in normal and fibrotic liver samples of mice using immunohistochemical (IHC) analysis. A low level of DGKG in the normal livers was seen surrounding portal veins. In contrast, the fibrotic livers exhibited significantly increased DGKG expression in the areas surrounding egg granuloma damage boundaries and blood vessels (Fig. 4b). To identify the cell types in which DGKG is expressed around egg granulomas during fibrogenesis, we co-localized DGKG with cellular markers in mice fibrosis liver using immunofluorescent co-staining. These cell markers include hepatocytes (Albumin), hepatic stellate cells (α-SMA and GFAP), macrophages (F4/80), epithelial cells (CK19) and leukocytes (CD45) (Supplementary Fig. 3). We found that DGKG co-localized with albumin-positive signals in hepatocyte around granulomas and blood vessels in infected livers, with some co-staining near blood vessels in normal liver tissue (Fig. 4c, d and Supplementary Fig. 3).

### DGKG facilitates the pathological progression and hepatic fibrosis after *S. japonicum* infection

To test whether DGKG regulates liver fibrogenesis, wild-type (WT) mice were injected with HBAAV2/8-m-*Dgkg* targeting the liver to overexpress *Dgkg* (AAV-*Dgkg*), while mice injected with empty AAV vector served as controls (AAV-Ctrl). The *in vivo* imaging system confirmed the successful transduction of AAV into the liver (Supplementary Fig. 4a, b). Western blot results showed a significant upregulation of DGKG expression in the fourth week after AAV transduction compared to the AAV-Ctrl group (Supplementary Fig. 4c). To ensure sustained high expression of DGKG during the fibrotic process, mice were injected with AAV via tail vein two weeks before *S. japonicum* infection (Fig. 5a). Overexpression of the *Dgkg* gene led to greater weight loss and more severe hepatic fibrosis in *S. japonicum*-infected mice for seven weeks, as indicated by western blotting of a-SMA (Supplementary Fig. 5a, b). We further used *Dgkg* knockout (*Dgkg*-KO, homozygous, Supplementary Fig. 6a, b) mice infected with *S. japonicum*. Surprisingly, compared to WT, *Dgkg*-KO mice exhibited less weight loss and dramatically attenuated fibrosis as measured by western blotting of a-SMA (Supplementary Fig. 6c, d).

**Fig. 5.**
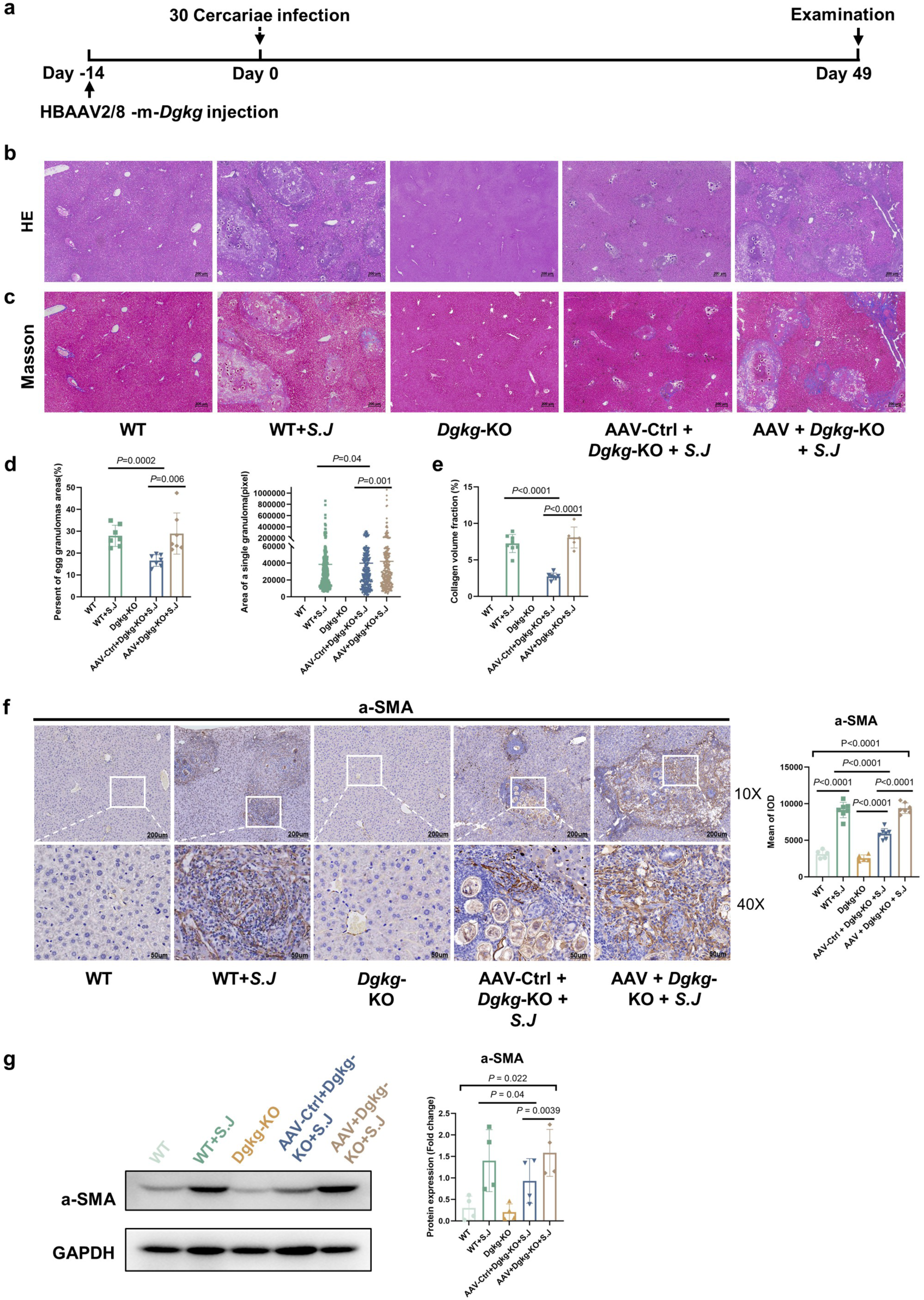
DGKG aggravates the pathological progression of schistosomiasis. (a) Time schedule for parasite infection, tail vein injection of AAV vectors, and sample examination. (b) H&E staining of liver sections. (c) Fibrosis was observed using liver sections stained with Masson’s trichrome. (d) The percentage of granulomatous area and the area of a single granuloma were measured from H&E sections using Image-Pro Plus 6.0 software (n = 7 per group). (e) The integrated optical density (IOD) of collagen volume fraction was analyzed by Image-Pro Plus 6.0 (n = 7 per group). (f) Expression of α-SMA in mice livers was analyzed by IHC (n = 5-8 per group) and the mean of the IOD was analyzed by Image-Pro Plus 6.0. (g) Protein expression of α-SMA in mice livers was analyzed by western blotting (n = 4 per group). Bars and error bars represent the mean ± SD, respectively. Statistical analysis was performed using paired Student’s T-test (pair samples with similar egg counts per gram of liver tissue).

Then we injected HBAAV2/8 to rescue and overexpress *Dgkg* in knockout mice, with mice injected with the empty AAV vector as a control. To summarize, we designed five experimental groups: (1) *Dgkg*-KO mice injected with AAV combined with *S. japonicum* infection (AAV + *Dgkg*-KO + *S.J*), (2) *Dgkg*-KO mice injected with AAV-Ctrl combined with *S. japonicum* infection (AAV-Ctrl + *Dgkg*-KO + *S.J*), (3) *Dgkg*-KO mice without infection (*Dgkg*-KO), (4) wild-type mice combined with *S. japonicum* infection (WT + *S.J*), and (5) wild-type mice without infection (WT). We monitored the body weight changes of mice during the progression of schistosomiasis (Supplementary Fig. 7a). We found that the body weight of mice overexpressing *Dgkg* was significantly lower than that of the *Dgkg*-knockout group after seven weeks of *S. japonicum* infection (*P* = 0.037). Three weeks after infection, the infected mice began to lose weight due to the deposition of *S. japonicum* eggs in the liver, especially those in AAV + *Dgkg*-KO + *S.J* group, which had the fastest weight loss after two weeks of infection because of DGKG overexpression. In contrast, the AAV-Ctrl + *Dgkg*-KO + *S.J* group showed the slowest weight loss. There were no significant differences in egg burden among these infected groups, indicating the transfection of AAV did not affect parasite reproduction (Supplementary Fig. 7b). Our *in vivo* experimental modeling time ended at the seventh week of *S. japonicum* infection because mice in group AAV-*Dgkg* began to die and exhibited adverse symptoms such as severe weight loss being underweight. According to this result, *DGKG* is related to body weight or BMI, as reported in the GWAS catalog, suggesting that *DGKG* may be involved in body metabolism.

Interestingly, H&E staining of liver sections showed that the percent of egg granuloma areas, and the area of a single granuloma were significantly shrunk (44.1%) in the AAV-Ctrl + *Dgkg*-KO + *S.J* group compared to those in the control group, (Fig. 5b, d). Furthermore, Masson’s trichrome staining showed a significant reduction in collagen deposition in the livers of mice in the AAV-Ctrl + *Dgkg*-KO + *S.J* group (Fig. 5c, e). We examined the α-SMA expression in the liver tissues using western blotting and IHC. As expected, α-SMA expression was significantly decreased (33.5%) in the AAV-Ctrl + *Dgkg*-KO + *S.J* group compared to the AAV + *Dgkg*-KO + *S.J* group (*P*=0.002), exhibiting dramatically attenuated fibrosis (Fig. 5f, g). Additionally, *Dgkg* overexpression in mice infected with *S. japonicum* led to marked hepatosplenomegaly (Supplementary Fig. 7c). The spleen index was significantly decreased in the AAV-Ctrl + *Dgkg*-KO + *S.J* group compared with the WT + *S.J* group (*P* = 0.029) and the AAV + *Dgkg*-KO + *S.J* group (*P* = 0.033). qRT-PCR results showed that *Dgkg* overexpression significantly increased the levels (*P*<0.05) of inflammatory factors (TNF-a, IL10, IL6) in the infected liver tissues (Supplementary Fig. 7d). These findings establish a causal role of DGKG in schistosomiasis-induced hepatic fibrosis, demonstrating that DGKG promoted the pathological progression, enhanced inflammatory responses, and exacerbated hepatic fibrogenesis following *S. japonicum* infection.

### DGKG influences lipid signaling, energy metabolism, cell cycle, and immune responses

We performed RNA-seq and differential gene expression analysis on liver tissues from five groups of mice (threshold: *P*-adj ≤ 0.05, log_2_FC ≥ |1|). In the comparison between *Dgkg*-KO group and WT group, a total of 1,745 differentially expressed genes (DEGs) were identified, comprising 828 upregulated and 917 downregulated genes (Fig. 6a). Notably, GO and KEGG enrichment analyses revealed that upregulated genes were predominantly involved in lipid metabolism and energy metabolism pathways, including the PPAR signaling pathway, fatty acid synthesis and degradation, glyceride metabolism, peroxisomal functions, and retinol metabolism (Fig. 6b). Conversely, downregulated genes were primarily involved in immune responses, such as leukocyte migration, chemotaxis, and T cell activation (Fig. 6c). In the comparison between the *Dgkg*-KO + *S.J* and WT + *S.J* groups, 1,806 DEGs were identified (828 upregulated, 978 downregulated; *P*-adj*_Dgkg_* = 0.01, log_2_FC*_Dgkg_* = 1.77; Fig. 6d). Enrichment analysis indicated that, in addition to lipid metabolism and energy metabolism, upregulated genes were also linked to cytochrome P450-mediated metabolism of xenobiotics (Fig. 6e). Downregulated genes, besides being involved in immune responses, were also associated with B cell receptor signaling, cytokine-cytokine receptor interactions, cell cycle regulation, and extracellular matrix (ECM)-receptor interactions (Fig. 6f). As expected, the DEG enrichment patterns for the AAV + *Dgkg*-KO + *S.J* vs. AAV-Ctrl + *Dgkg*-KO + *S.J* comparison were consistent with those observed in the WT + *S.J* and *Dgkg*-KO + *S.J* comparison (Supplementary Fig. 8); thus, no redundant details are provided here. These findings suggest that DGKG may modulate hepatic fibrosis progression by regulating lipid metabolism and immune responses.

**Fig. 6.**
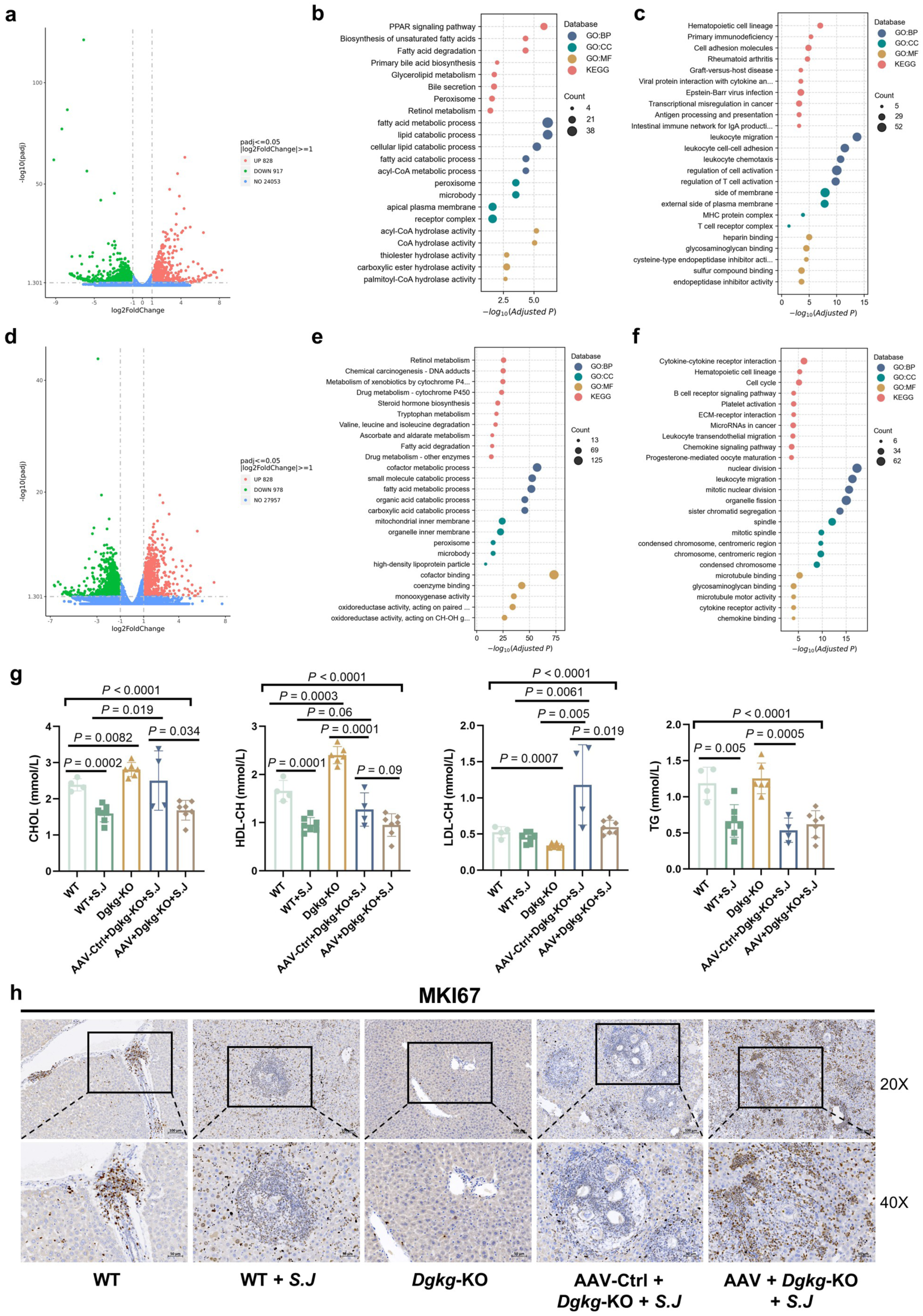
DGKG influences lipid signaling, energy metabolism, cell cycle, and immune responses. (a) Volcano plots of differential expression of genes between *Dgkg*-KO vs. WT groups. (b, c) Bubble plots of the GO terms and KEGG pathways of (b) up-regulated and (c) down-regulated genes between *Dgkg*-KO vs. WT groups, respectively. (d) Volcano plots of differential expression of genes between AAV-Ctrl+*Dgkg*-KO+*S.J* vs. WT+*S.J* groups. (e, f) Bubble plots of the GO terms and KEGG pathways of (e) up-regulated and (f) down-regulated genes between AAV-Ctrl+*Dgkg*-KO+*S.J* vs. WT+*S.J* groups, respectively. (g) Serum levels of the liver function test parameters in mice. (h) Expression of MKI67 in mice livers was analyzed by IHC. Bars and error bars represent the mean ± SD. Statistical analysis was performed using the Student’s T-test.

In addition, *Dgkg* knockout mice exhibited significantly elevated levels of circulating cholesterol (CHOL) and high-density lipoprotein cholesterol (HDL-CH) compared to wild-type controls, irrespective of *S. japonicum* infection status (Fig. 6g). Conversely, *Dgkg* overexpression resulted in a marked reduction in CHOL levels (*P* = 0.034) relative to the AAV-Ctrl + *Dgkg*-KO + *S.J* group. There is no significant difference in low-density cholesterol lipoprotein (LDL-CH) levels observed between the infected and uninfected wild-type mice. But in *Dgkg* knockout mice, LDL-CH level is significantly increased after *S. japonicum* infection (*P* = 0.0061), and significantly decreased compared to uninfected wild-type mice (*P* = 0.0007). Triglyceride (TG) levels remained unaffected by DGKG expression. Immunohistochemical analysis of MKI67 further revealed that *Dgkg* overexpression significantly promotes liver cell proliferation (Fig. 6h), indicating a potential role for DGKG in cell cycle regulation. Moreover, pronounced hepatic glycogen accumulation was observed in the AAV + *Dgkg*-KO + *S.J* group, whereas the glycogen storage in the *Dgkg* knockout mice was lower than that in wild-type controls (Supplementary Fig. 9). These findings suggest that DGKG promotes pathological progression and hepatic fibrosis, and modulates liver inflammation following *S. japonicum* infection. Moreover, elevated DGKG expression may affect lipid and energy metabolism, disturb cell cycle regulation, and aggravate liver injury.

## DISCUSSION

Our genome-wide association study identifies eight novel loci associated with *S. japonicum*-induced hepatic fibrosis, including the genome-wide significant locus at 16p13 (rs73575170, *P* = 3.9×10^-8^). Through integrative mapping (positional, eQTL, chromatin interaction, and ECS), we mapped 262 candidate genes, including the prioritized *DGKG*--a lipid metabolism hub--emerging as a regulator of lipid/energy metabolism and immune response. In vivo validation demonstrated that *Dgkg* knockout attenuates fibrosis progression, while overexpression exacerbates fibrotic and inflammatory markers (α-SMA↑, TNF-α↑, *P* < 0.05), establishing its causal role in schistosomiasis pathology.

While prior studies focused on immunogenetic mechanisms (14, 15, 20, 50–54), our GWAS highlights metabolic susceptibility factors. Unlike viral hepatitis models, schistosomiasis fibrosis arises from chronic egg antigen-driven microenvironmental imbalance (2, 12), underscoring the need for etiology-specific therapeutic strategies. The absence of overlapping loci with other liver fibrotic diseases further emphasizes this divergence. Schistosome infection alters the hepatic tissue microenvironment, leading to an imbalance of intrahepatic immune-inflammatory microenvironments (55). Changes in liver inflammation processes, hepatic cell metabolic pathways, and other factors play a crucial role in shaping and regulating the local immune-inflammatory microenvironment in the liver. DGKG is expressed in hepatocytes surrounding granulomas and peri-portal vessels, linking metabolic reprogramming to local immune dysregulation. Previous studies in the UKBB have linked *DGKG* SNPs to metabolism, showing associations with BMI and lipoprotein cholesterol (41–43). Consistently, our in vivo experiments also demonstrated that *DGKG* influences body weight and cholesterol metabolism in mice.

The lead SNP rs6762330, located in the intron of the *DGKG* gene in the chromosome 3q27.3 region, exhibited a positive correlation with hepatic fibrosis (beta = 0.1234, with C as effect allele in our GWAS). Interestingly, an eQTL study in East Asian populations revealed that the T allele of this SNP (the non-effect allele in our GWAS) negatively correlated with *DGKG* expression in the blood (beta = -0.123) (56). Thus, it can be inferred that the C allele of this SNP may upregulate *DGKG* expression, exacerbating the degree of fibrosis. Remarkably, our experimental findings also support this inference, as we observed a positive correlation between *DGKG* expression and the hepatic fibrosis progression at different time points in the livers of mice infected with *S. japonicum* (*r* = 0.816, *P* < 0.0001, Fig. 4a).

The gene *DGKG* encodes diacylglycerol kinase gamma (DGKG), an enzyme that phosphorylates diacylglycerol (DAG) to produce phosphatidic acid (PA) (57). During *S. japonicum* infection, deposition of parasite eggs in the vasculature triggers the release of egg-derived antigens, which recruit immune cells and promote granuloma formation(2). Within this inflammatory and hypoxic microenvironment, surrounding hepatocytes undergo metabolic reprogramming (58). PA not only acts as a lipid second messenger but also provides an essential precursor for Glycerol-3-phosphate (Gro3P)-derived lipid biosynthesis, bridging lipid signaling with energy metabolic adaptation (59, 60). Gro3P shuttle mediates the transfer of glycolysis-derived NADH into mitochondrial oxidative phosphorylation for ATP production under hypoxic conditions (61). Moreover, Gro3P functions as a central metabolic node linking energy metabolism, gluconeogenesis, and lipid biosynthesis (62, 63). Mechanistic studies have indicated that DGKG can influence glucose uptake, lactate, and ATP generation by affecting glucose transporter-1 and MAPK (64). Consistently, DGKG deficiency attenuated inflammation and fibrosis in *S. japonicum*-infected mice, accompanied by enhanced hepatic lipid and energy metabolism, whereas DGKG overexpression produced the opposite effects. These findings suggest that DGKG may promote schistosomiasis-induced liver fibrosis by orchestrating lipid-immune crosstalk (Fig.7).

**Fig. 7.**
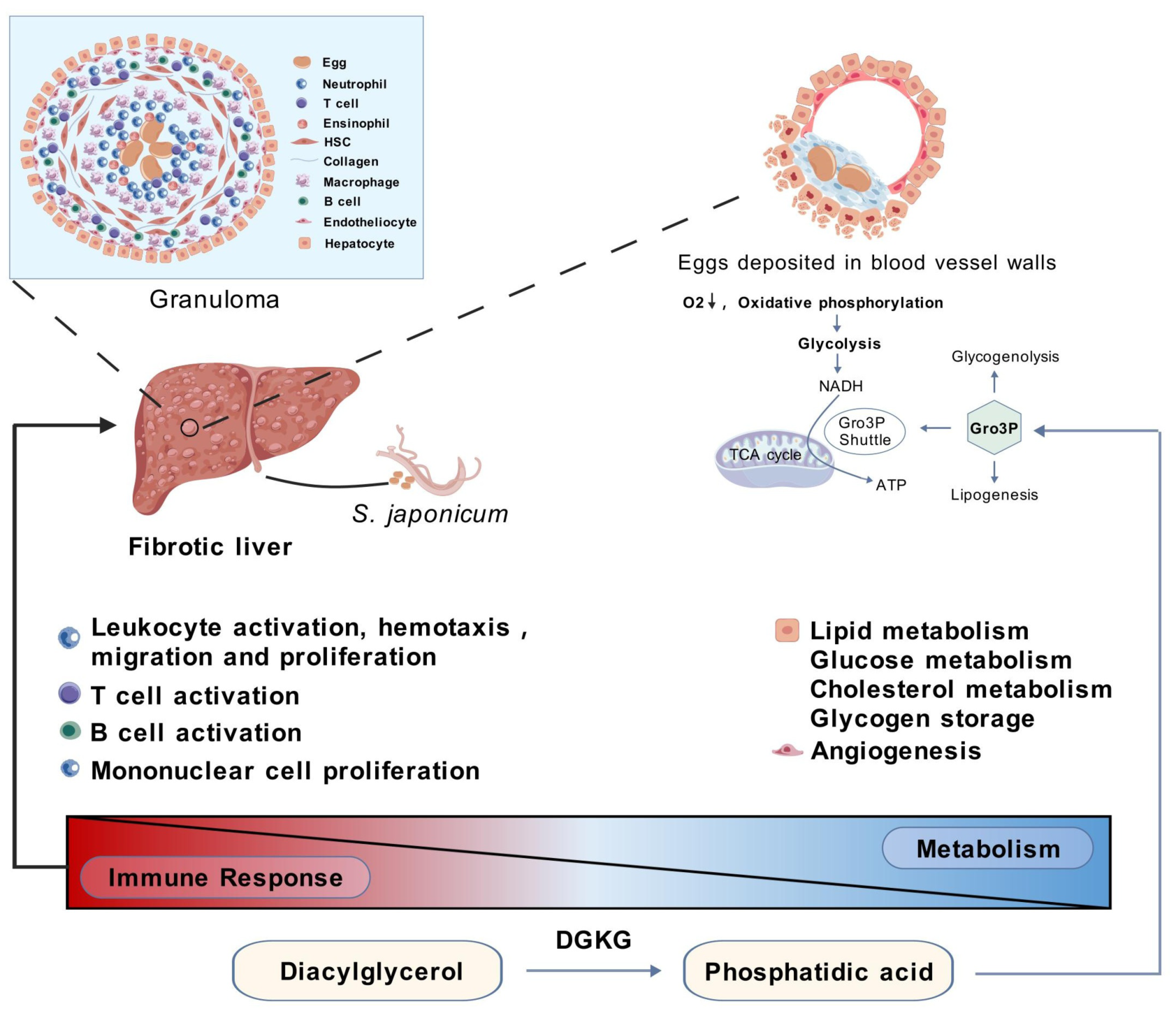
A potential mechanism by which DGKG regulates schistosomiasis-induced liver fibrosis. Rectangle colors from blue to red indicate low to high DGKG expression. Triangles labeled “Immune response” and “Metabolism” represent the upregulation of immune-related pathways in high DGKG expression and metabolism-related pathways in low DGKG expression, respectively. Created with BioGDP.com (65).

Schistosome infection induces profound metabolic and immune shifts, yet schistosomiasis fibrosis lacks genetic overlap with other fibrotic diseases, underscoring its distinct etiology. This suggests that metabolic regulation plays a unique role in schistosomiasis hepatic fibrosis, rather than being a secondary consequence of tissue damage. Importantly, our GWAS identifies *DGKG* as a novel fibrosis susceptibility gene in an infectious disease setting, broadening our understanding of host genetic factors in parasite-induced fibrosis.

As uninfected individuals in the general population still risk developing severe schistosomiasis, comparing the contribution of SNPs between uninfected subjects and patients with severe schistosomiasis may limit the ability to obtain positive results. Therefore, all samples were obtained from *S. japonicum*-infected patients to avoid type II errors (false negatives). Nevertheless, our analysis has certain limitations. First, while our findings support *DGKG* as a regulator of fibrosis, independent validation in other schistosomiasis cohorts is needed. Second, our sample size, though substantial for a neglected tropical disease study, remains modest compared to other GWAS. However, we employed stringent quality control, comprehensive multi-omic analyses, and knock-out mice experiments to enhance result reliability.

Overall, we provide the first genome-wide evidence linking *DGKG* to schistosomiasis hepatic fibrosis and uncover its role in lipid-immune crosstalk. These findings advance our understanding of the metabolic genetic basis of fibrosis and offer potential therapeutic targets for schistosomiasis and related fibrotic disorders.

## MATERIALS AND METHODS

### Participants and data collection

To discover new susceptibility sites for schistosomiasis hepatic fibrosis, 984 patients with a history of schistosomiasis japonica were recruited through the epidemiological survey in nine counties, randomly selected from those within the scope of medical assistance for late-stage schistosomiasis japonica, around Poyang Lake in Jiangxi Province of China from 2015 to 2017. According to the “Diagnostic Criteria for Schistosomiasis” issued by the Ministry of Health of the People’s Republic of China in 2006 and “The Prevention and Control of Schistosomiasis and Soil-Transmitted Helminthiasis” reported by the WHO, participants were selected by professional laboratory and pathologists after evaluation. The inclusion criteria of the study subjects included: (1) a history of long-term or repeated exposure to infected water; (2) a clear history of treatment for schistosomiasis japonica, or fecal examination found worm eggs or miracidia, or rectal biopsy found schistosomiasis eggs, or positive serum immunological test for long-term; (3) Exclude the patients who self-reported a history of schistosomiasis, but were negative in serum immunological examination and had no information of B-ultrasound liver fibrosis. For schistosomiasis patients who met the above criteria, ten milliliters of anticoagulated peripheral blood samples were collected on an empty stomach for breakfast, and the serum and blood cells were separated by centrifugation (4000rpm, 10 minutes) on the investigation site. Serum samples were used for biochemical index detection, and blood cells were used to extract genomic DNA for SNP genotyping. Samples were stored in a -80°C freezer. The on-site investigation was completed with the support of the Jiangxi Provincial Institute of Parasitic Disease Control, and all participants signed informed consent.

Biological information was collected at the local schistosomiasis control station *via* face-to-face questionnaire interviews and physical examination. The physical examinations such as blood pressure, height, weight, and waist circumference of the research subjects were repeated three times, and the average value was taken. The questionnaire contained sociodemographic characteristics (age, education level, occupation, and income level, e.g.), medical history of schistosomiasis, history of contact with infected water contact, and life behavior. The portal vein diameter, ascites, and related imaging markers were measured by abdominal ultrasonography. The detection of biochemical indicators was completed by KingMed Diagnostics (Guangzhou, China).

### Animal and ethics statement

The studies involving human participants were reviewed and approved by the Ethics Committees of Fudan University (IRB#2016-TYSQ-03-17). The patients/participants provided written informed consent to participate in this study. Animals were cared for according to the guidelines developed by the China Council on Animal Care, and all animal experiments were performed according to the procedures approved by the Human Research Ethics Committee of Sun Yat-sen University (No·2016-104), SYXK (Guangdong) 2015-0107 and SYSU-IACUC-2023-B1013.

Wild-type male C57BL/6 mice were purchased from Guangdong Medical Laboratory Animal Centre. *Dgkg* knock-out mice were purchased from GemPharmatech Co., Ltd. (Nanjing, China). Genomic DNA was extracted from the *Dgkg* KO mice and PCR amplified using the forward (F) and reverse (R) primers: PCR1 (F: 5’-AGGAAAGAGGTCAAAAGGAACCAGG-3’, R: 5’-GCCTAATTTTCAAGCAGAAATGTCAA-3’), PCR2 (F: 5’-TGGCCTGAGCTTCATTAGCAGTTT-3’, R: 5’-GATGGAGTCTTATAGGACAACACGGG-3’). Genotyping results showed homozygous (Supplementary Fig. 6a, b). All studies were performed in male mice. Mice were kept in a standard 12 h light-dark cycle under the specific-pathogen-free conditions and allowed free access to water and food.

To induce infection, we exposed percutaneously to *S. japonicum* cercariae that were shed from lab-infected snails (*Oncomelania hupensis*), obtained from the National Institute of Parasitic Disease, Chinese Centre for Disease Control and Prevention. All animal experiments were approved by the Sun Yat-sen University Committee for Animal Research and conformed to the Guidelines for the Care and Use of Laboratory Animals of the National Institute of Health in China.

### Definition

Based on the *Diagnostic Criteria for Schistosomiasis* established by the Department of Disease Control of the Chinese Ministry of Health in 2006, this study has implemented more rigorous definition standards for chronic and advanced-stage schistosomiasis. Chronic schistosomiasis: (1) Meets the diagnostic criteria for *S. japonicum*; (2) Abdominal B-ultrasound reveals hepatic fibrosis grade 0-1, with a portal vein diameter below 12mm; (3) Absence of ascites, hepatosplenomegaly, and other symptoms. Advanced schistosomiasis: (1) Meets the diagnostic criteria for *S. japonicum*; (2) Abdominal B-ultrasound reveals hepatic fibrosis grade 3, with a portal vein diameter ≥13mm; (3) Presence of clinical manifestations such as ascites, hepatosplenomegaly, gastroesophagoduodenal varices, or upper gastrointestinal bleeding.

The degree of hepatic fibrosis was assessed by a professional physician using abdominal ultrasonography. According to the *recommendations of the WHO Cairo Working Group in 1992*, the *Schistosomiasis Prevention and Control Manual* issued by the Chinese Ministry of Health Disease Control Department, and the *Technical Guidelines for Schistosomiasis Prevention and Control*, the liver was examined comprehensively in a supine and fasting position with quiet respiration using routine liver sections such as the transverse section at the level of the xiphoid process and the oblique section beneath the right ribs. The echo intensity of the liver was compared to that of the renal parenchyma, and the hepatic fibrosis was graded into four levels: Grade 0 indicates no fibrosis; Grade 1 indicates mild fibrosis with increased coarse echogenicity and granularity; Grade 2 indicates moderate fibrosis with uneven echogenicity, diffuse fine-meshed echoes, slightly thickened liver vessel walls, and normal hepatic vasculature; Grade 3 indicates severe fibrosis with highly uneven echogenicity, diffuse coarse-meshed echoes, markedly thickened portal vein walls, narrowed liver vessel lumen, unclear display of intrahepatic vessels, and reduced liver volume.

### Genotyping, quality control, and imputation

Based on an automated nucleic acid purification platform (BioTeke, AU1001, Beijing, China), genomic DNA was extracted from the 200 ml of frozen blood cells using the magnetic bead-based method for concentrating DNA (The Genomic DNA Magnetic Beads Kit, AU18016, BioTeke, Beijing, China). Then, the Illumina Infinium Asian Screening Array was used to genotype 659,184 SNPs. After stringent quality filtering, 497,495 SNPs from 900 samples were ultimately retained. Next, to extend the coverage to the genomic region, autosomal SNPs that passed strict quality checks were used to impute genotypes of SNPs across the chromosomes for all subjects, and 6,237,938 SNPs were genotyped. Finally, after quality controls and relatedness removal, a total of 4,523,335 remained, representing 637 unrelated individuals.

We performed strict quality control on samples and SNPs to guarantee robust association analyses. Briefly, samples were excluded if they (a) had an overall genotyping rate of <90%, (b) showed unexpected duplicates or relatives (PI_HAT > 0.05 calculated by KING (66)), or (c) were identified as outliers. PCA was used to detect the outliers using Genome-wide Complex Trait Analysis software (GCTA; version 1.92.2). Twenty principal components were calculated, and the first four ancestry principal components explained 60% variation. In addition, SNPs were excluded if (a) the call rate was <90%; (b) minor allele frequency (MAF) was <0.05; (c) did not map to autosomal chromosomes; and (d) if the P-value was less than 1.0 × 10^-5^ in Hardy-Weinberg equilibrium tests.

Following the prephasing of genotypes with Shape-IT (67) v2, we imputed genetic markers from the 1000 Genomes Project reference panel (phase III integrated release; http://1000genomes.org) using IMPUTE2 (68) v2.3.0. We used the information *r^2^* from IMPUTE2 as a QC parameter, which estimates the correlation between imputed and true genotypes. Genetic markers with an imputation *r^2^* < 0.4 and quality score < 0.4 were removed.

### GWAS analysis

We used PLINK (69) v1.9 (www.cog-genomics.org/plink/1.9/) to analyze the associations between genome-wide SNPs and schistosomiasis hepatic fibrosis with a linear regression model. For the selection of PCA covariates, we used EIGENSTRAT (70) to calculate the number of significant principal components (PCs) and selected the first seven significant PCs based on a *P* value of 0.05. We used non-genetic factors (characteristics in Table 1) as independent variables to construct linear regression models. Then, stepwise regression was used to screen the optimal set of covariates. We modeled data from each genetic marker using additive dosages to account for imputation uncertainty. The effective number of independent SNPs was calculated in the Genetic Type 1 Error Calculator (GEC) (71) (http://pmglab.top/gec/). The effective number of independent tests for all the 3,317,263 valid variants is 303,884.86 (9.16%). The p-value cutoff by Bonferroni correction for family-wise error rate 0.05 is 1.65E-07. The genome-wide significant *P* value cutoff by Bonferroni correction was approximately 1.65 × 10^−7^, and the suggestive *P* value was 1 × 10^−5^.

### Genomic risk locus definition

Genomic risk loci were defined using FUMA (36), an online platform for functional mapping and annotation of genetic variants. We first identified significant independent SNPs with a suggested significant two-tailed *P* value (*P* < 1 × 10^−5^) and which were independent of each other at *r^2^* < 0.6. Lead SNPs or index SNPs further represented these SNPs, a subset of the significant independent SNPs, with the most significant *P* value in approximate linkage equilibrium. Next, we defined independent genomic risk loci by merging any physically overlapping lead SNPs (LD blocks < 250 kb apart). The borders of the genomic risk loci were determined by identifying all SNPs in LD (*r^2^* ≥ 0.6) with one of the significant independent SNPs in the locus; the region containing all of these candidate SNPs was considered a single independent genomic risk locus. All LD information was calculated using the 1000 genome phase III EAS genotype data as a reference.

### Functional annotation of SNPs

Functional annotation of the SNPs implicated in this study was performed using FUMA. We selected all candidate SNPs in genomic risk loci with an *r^2^* ≥ 0.6 with one significant independent SNPs, a *P*-value 1 × 10^−5^, and an MAF > 0.05. Predicted functions of these SNPs were obtained by matching their chromosomes, base-pair positions, and reference and alternate alleles to databases containing known functional annotations, including CADD and RegulomeDB. CADD scores predict how harmful the effect of an SNP is likely to be for non-coding gene/protein structure/function, with higher scores referring to higher deleteriousness. A CADD score above 12.37 is the threshold for potential pathogenicity. The RegulomeDB score is a categorical score based on eQTLs and chromatin marks, ranging from 1a to 7, with lower scores indicating an increased likelihood of having a regulatory function. The chromatin state represents the accessibility of genomic regions (every 200 bp). A lower state means higher accessibility, with states 1–7 referring to open chromatin states. We annotated the minimum chromatin state across tissues to SNPs.

### Gene mapping

Significant loci obtained by GWAS were mapped to genes in FUMA using three strategies:

1. Positional mapping of SNPs to genes based on physical distance (within a 10 kb extension at the transcription start and end sites) from known protein-coding genes in the human reference assembly (GRCh37/hg19).
2. eQTL mapping of SNPs to genes with which they showed a significant eQTL association (i.e., allelic variation at the SNP is associated with the expression level of that gene). eQTL mapping uses information from 54 tissue types in three data repositories (GTEx, Blood eQTL browser, BIOS QTL browser) and is based on cis-eQTLs that can map SNPs to genes up to 1 Mb away. We used a false discovery rate (FDR) of 0.05 to define significant eQTL associations.
3. CI mapping of SNPs to genes when there are DNA–DNA interactions between the SNP and gene regions. CI mapping can involve long-range interactions, as it does not have a distance boundary, and an interacting region can span multiple genes. We used an FDR of 1 × 10−5 to define significant interactions, based on previous recommendations modified to account for the differences in the cell lines used here.

### Gene-based analysis

Gene-based association analysis was conducted using the ECS method (22) from the KGG platform (https://pmglab.top/kgg), with a total of 25,216 genes included in the analysis. Genotype data from East Asian individuals in the 1000 Genomes Project (72) were used for linkage disequilibrium (LD) calculations. In comparison with gene mapping (the Venn plot), a gene-level p-value threshold was set at 1E-4 to prioritize genes in the moderate GWAS sample.

### Estimating associated cell types

We employed the Phenotype-Cell-Gene Association (PCGA) analysis platform (22, 44, 47) to estimate the associated cell types. The LD calculations used genotypes from East Asian individuals in the 1000 Genomes Project. A gene-level p-value threshold of 0.05 was set, with a maximum of 500 genes. For cell type detection, we selected 82 cell clusters from human liver single-cell data and applied Bonferroni correction for multiple testing of *P*-values.

### KOBAS enrichment

The GO classification and KEGG pathway enrichment were conducted using KOBAS (73) 3.0 with hypergeometric test / Fisher’s exact test (*P* < 0.05). The input gene list is the gene whose ECS *P-*value is less than 0.01.

### qRT-PCR and western blotting

The expression of target mRNA was determined using the SYBR Green Master Mix kit (Takara, Japan). GAPDH was used as an internal control, and the fold change was calculated by the 2^−ΔΔCT^ method. For additional details, see supplementary information.

Liver tissues were homogenized with RIPA lysis buffer with freshly added protease and phosphatase inhibitors (Thermo Fisher Scientific, USA). The resulting total lysates were assayed for protein concentration using the BCA protein assay kit. Lysates were subjected to 10% sodium dodecyl-polyacrylamide gel electrophoresis and transferred to a polyvinylidene fluoride blotting membrane. The membranes were then immunoblotted with antibodies. For additional details, see supplementary information.

Results are expressed as mean ± SD values. The data of both groups were analyzed using the unpaired two-sample t-test. Multiple comparisons between more than two groups were analyzed by one-way ANOVA or Kruskal-Wallis (non-parametric) test. The value of *P* < 0.05 was considered statistically significant.

### Recombinant adeno-associated virus (AAV) vectors and transduction

HBAAV2/8 -m-*Dgkg* viral particles were purchased from Hanbio Biotechnology Co., Ltd. Mice were injected via the tail vein with AAV (1.5 × 10^11^ v.g./ mouse) two weeks before infection with *S. japonicum*. The green fluorescent protein (GFP) expression of AAV in mice was observed using an in vivo imaging system and fluorescence microscopy.

### Histopathology and fibrosis measurement

Liver tissues from the infected mice were weighed and digested overnight in 4% potassium hydroxide, and released eggs were counted under a dissecting microscope. The index of liver and spleen was calculated based on the following formula: total weight of mouse liver or spleen (mg)/total weight of mouse body (g). The area of egg granulomas was measured based on H&E-stained sections using the Image-Pro Plus 6.0 software. The area of each granuloma in the section was measured. Fibrosis was observed using liver sections stained with Masson’s trichrome staining, and the integrated optical density (IOD) was analyzed by Image-Pro Plus 6.0. CHOL, HDL-CH, LDL-CH and TG levels were detected by KingMed Diagnostics (Guangzhou, China).

### Immunohistochemistry and immunofluorescence analysis

Liver tissues were fixed in 4% neutral-buffered formalin and embedded in paraffin. Sections were dewaxed and incubated with GFAP, α-SMA, Albumin, F4/80, CK19, CD45 and DGKG antibodies overnight at 4°C. The sections were then incubated with the indicated secondary antibodies. The sum of the IOD was analyzed by Image-Pro Plus 6.0.

## Data Availability

All data produced in the present study are available upon reasonable request to the authors.

## List of Supplementary Materials

Supplementary Figure 1 to 9 for multiple supplementary figures.

Supplementary Table 1 to 9 for multiple supplementary tables.

## Acknowledgments

We thank all the patients participating in this study. We thank the Jiangxi Provincial Institute of Parasitic Disease Control for its cooperation and support. This work was supported by grants from the National Natural Science Foundation of China (No. 32170637, No. 32300500), the Science and Technology Program of Guangzhou (No. 201803010116) and Guangdong project (No. 2017GC010644), China Postdoctoral Science Foundation (No. 2022M723666).

## Author contributions

Conceptualization, MZ, CX, WZ, XS, XW, and ML. Data curation, MZ and XW. Formal analysis, MZ and CX. Patient investigation and blood sample collection, MZ, LS, AN, BZ, JL, LZ, YL, YH, ZC, ZW, XW, and ML. Methodology: MZ, CX and ML. Visualization, MZ and CX. Wet experiments, MZ, CX, YH, YL, JS, LW, CC and XS. Writing - original draft, MZ. Writing - review and editing, all authors. Supervision, ZW, XW and ML. Funding acquisition, WZ, XS, and ML. All authors contributed to the article and approved the submitted version.

## Declaration of generative AI and AI-assisted technologies in the manuscript preparation process

During the preparation of this work, the author(s) used *ChatGPT* (*OpenAI*) and *Grok* (*xAI*) in order to improve the clarity, grammar, and readability of the manuscript text. After using these tools, the author(s) reviewed and edited the content as needed and take full responsibility for the content of the published article.

