## Supplementary Figures 1-9 for "GWAS Reveals Distinct Genetic Architecture of Schistosomiasis-Induced Hepatic Fibrosis with *DGKG* as a Key Mediator"

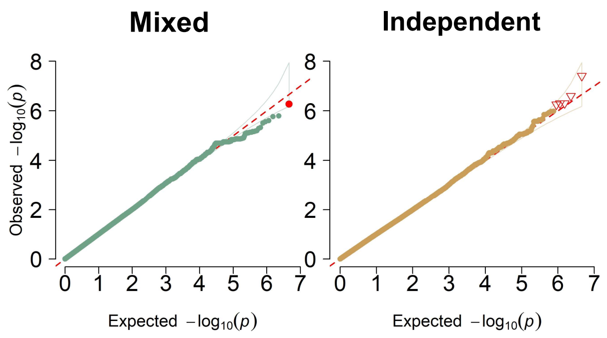


**Supplementary Fig.1 |** QQ-plot of Genome-wide association analysis of schistosomiasis hepatic fibrosis in mixed (N=900) and independent (N=637) sample cohorts.

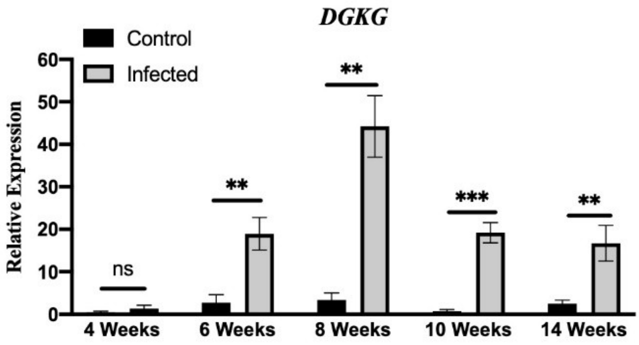


**Supplementary Fig. 2 | The expression of Dgkg in liver tissue of mice with different infection cycles was detected by qPCR.** ns means P＞0.05, ** P＜0.01, *** P＜0.000. Mean ± SEM.

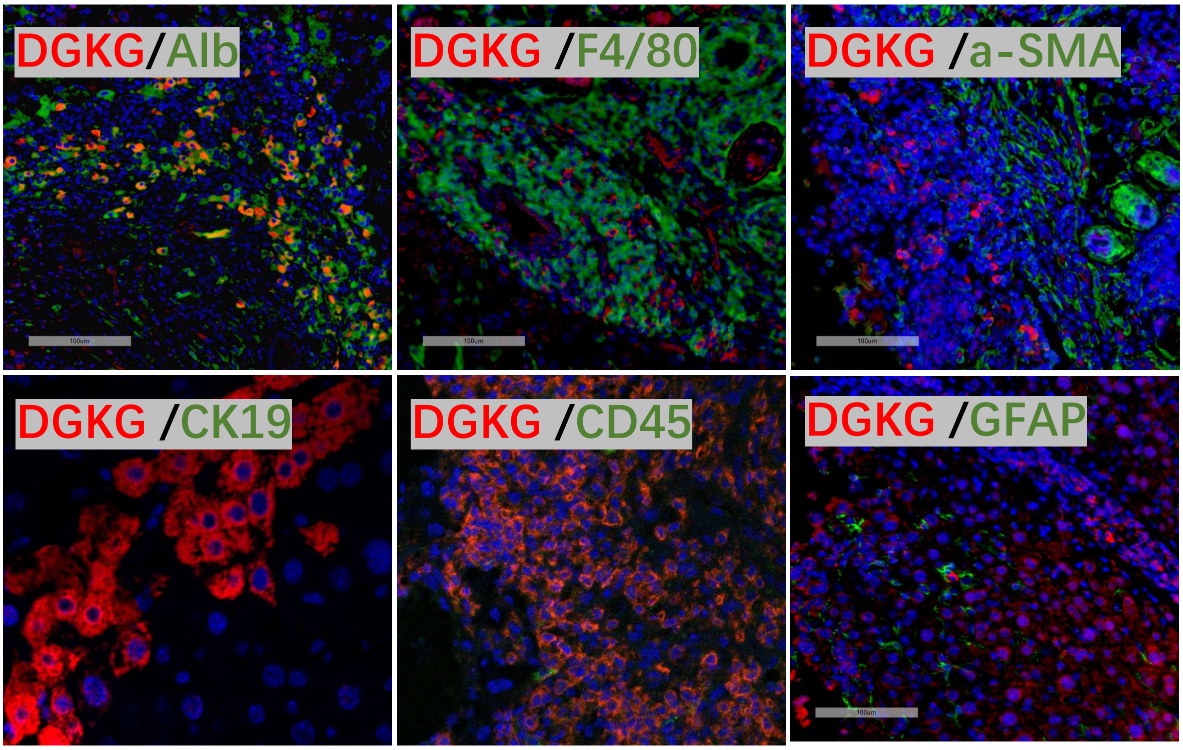


**Supplementary Fig. 3 | Immunofluorescence co-staining results of DGKG and cell markers.** The pictures show the liver tissue of mice infected with *S.japonicum*. DGKG co-localized with the hepatocyte marker Albumin.

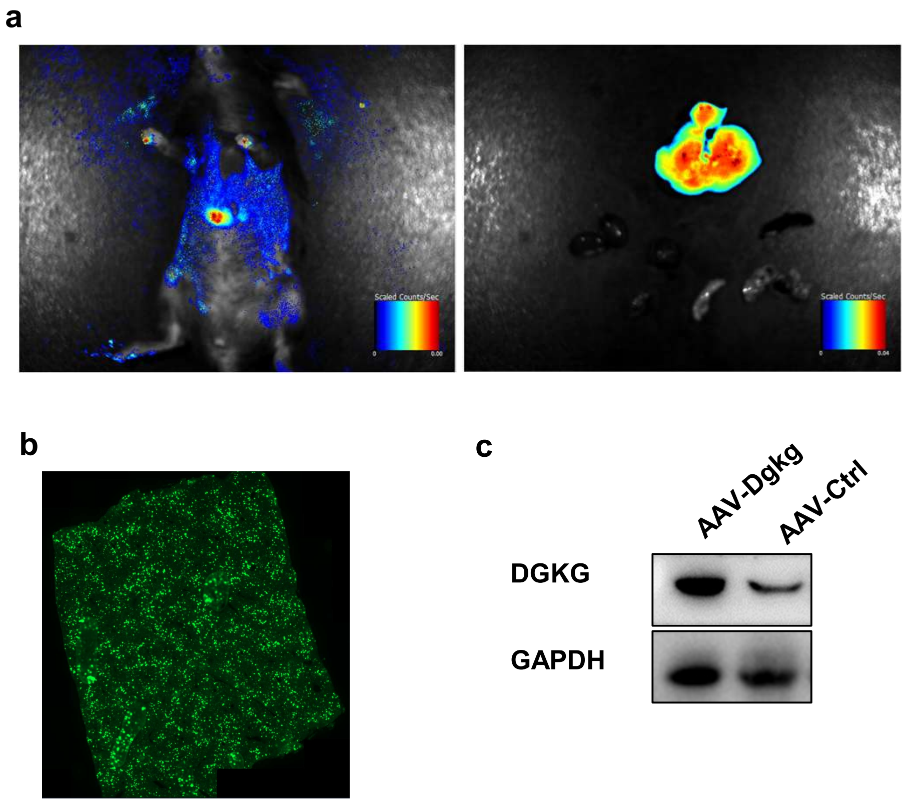


**Supplementary Fig. 4 | HBAAV2/8-m-Dgkg successfully transfected liver-targeted overexpressed adeno-associated virus.** (a) In vivo imaging and tissue imaging of mice. (b) Overall view of mouse liver infected with AAV. (c) Western blotting and IHC detection of the expression level of DGKG protein in mouse liver tissue after overexpressed AAV transfection for 4 weeks.

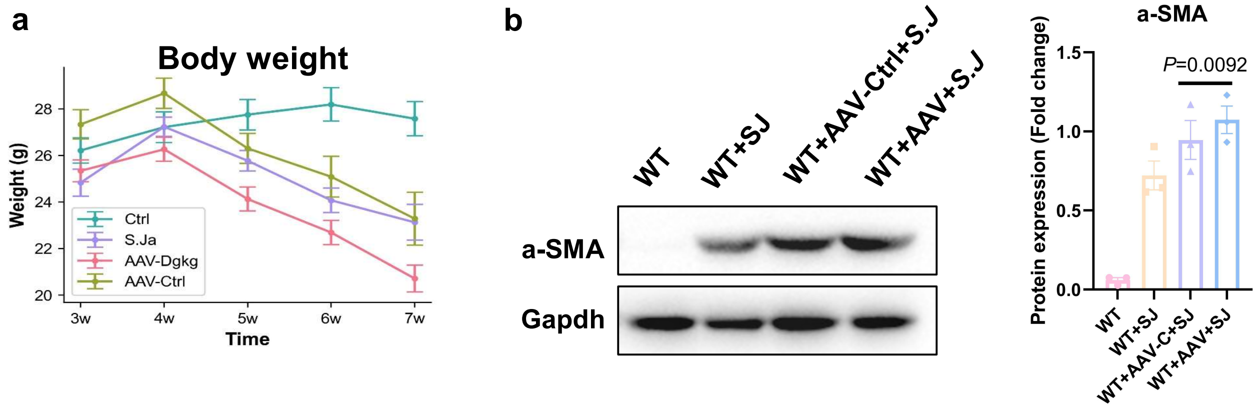


**Supplementary Fig. 5 | Infection experiment of wild-type mice injected with AAV overexpressing *Dgkg*.** (a) Body weight records of mice in each group during the 7 weeks infection. (b) Protein expression of α-SMA in mice livers were analyzed by western blotting (n = 3 per group). Bars and error bars represent the mean ± SD, respectively. Statistical analysis was performed using Student's T-test.

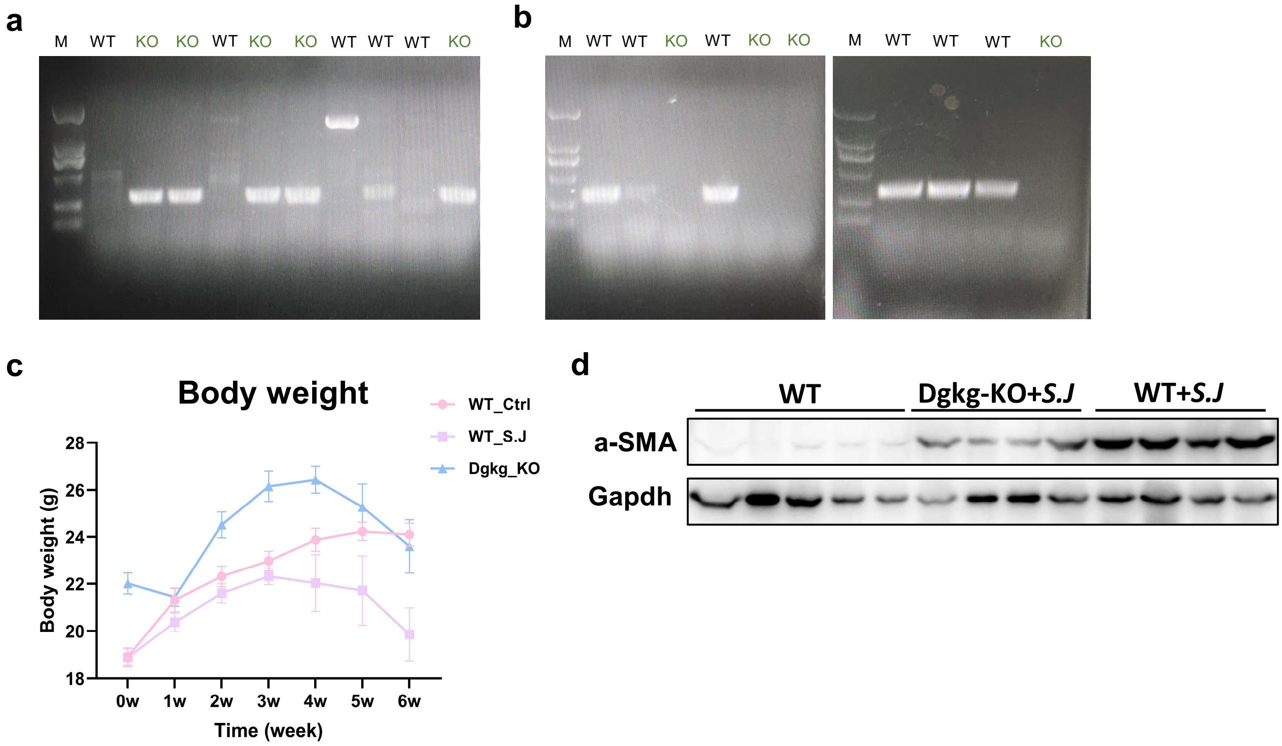


**Supplementary Fig. 6 | Infection experiment of *Dgkg* knockout mice.** (a) PCR reaction obtains a single KO band; (b) PCR reaction without product**.** (c) Body weight records of mice in each group during the 6 weeks infection. (d) Protein expression of α-SMA in mice livers were analyzed by western blotting (n = 4-5 per group).

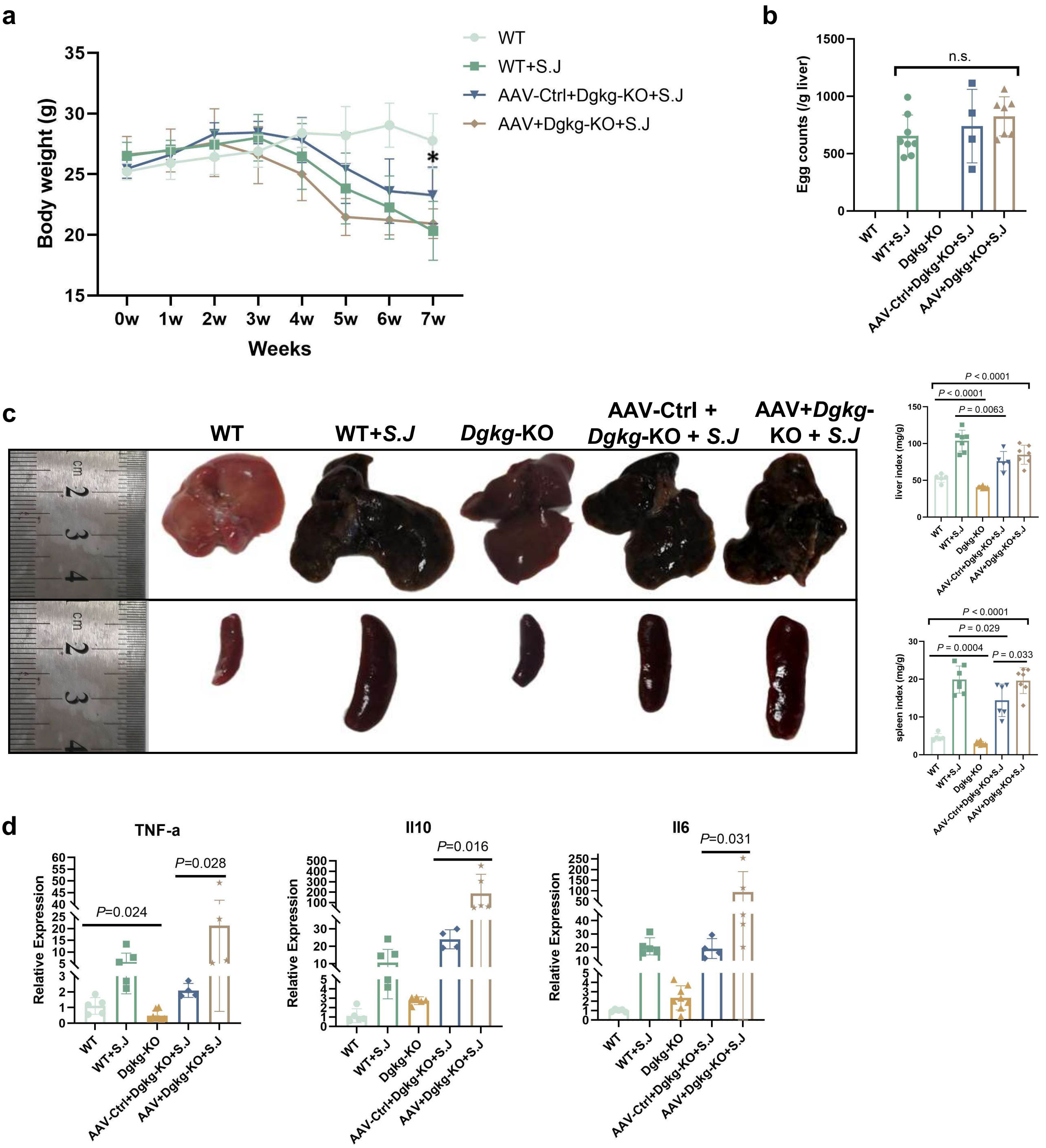


**Supplementary Fig. 7 | Body weight, liver, spleen, and inflammation levels of mice in each group 7 weeks after schistosomiasis infection.** (a) The body weight changes of mice during 0-7 weeks after *S.Japonicum* infection. * means student's t-test $P < 0.05$ for mice weight in AAV-Ctrl+*Dgkg*-KO+*S.J* group and AAV+*Dgkg*-KO+*S.J* group. (b) Egg counts per gram of liver tissue in the infected groups. (c) Upper: The macroscopic appearance of livers and liver index (n=5–8 per group). Below: The macroscopic appearance of spleens and spleen index (n=5–8 per group). (d) Expression of Inflammatory factors in liver tissues. Bars and error bars represent the mean ± SD, respectively. Statistical analysis was performed using a two-sided Student's T-test and one-way ANOVA Dunnett's multiple comparison test.

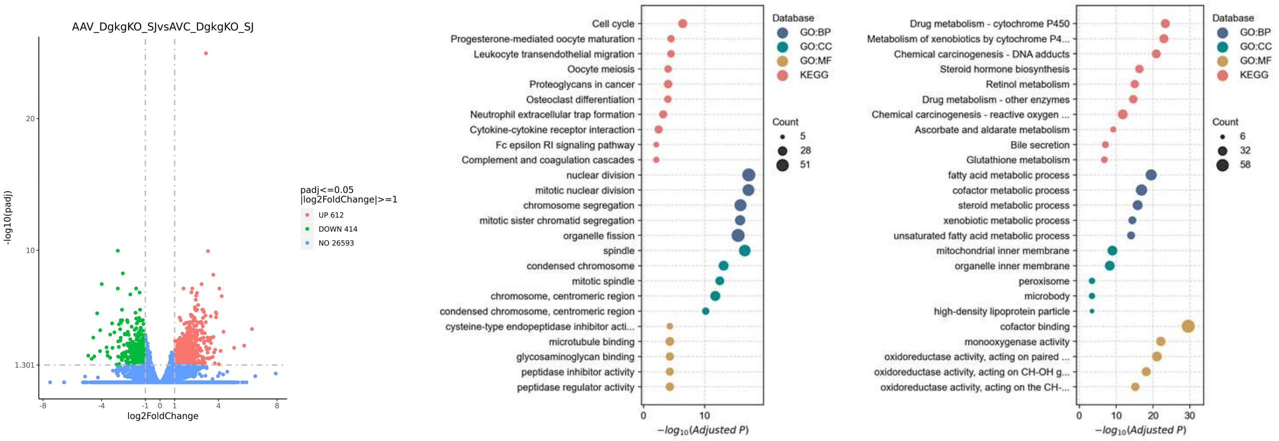


**Supplementary Fig. 8 | DGKG influences lipid signaling, energy metabolism, cell cycle and immune responses.** (a) Volcano plots of differential expression of genes between AAV+*Dgkg*-KO+*S.J* vs. AAV-Ctrl+*Dgkg*-KO+*S.J* groups. (b, c) Bubble plots of the GO terms and KEGG pathways of (b) up-regulated and (c) down-regulated genes between AAV+*Dgkg*-KO+*S.J* vs. AAV-Ctrl+*Dgkg*-KO+*S.J* groups, respectively.

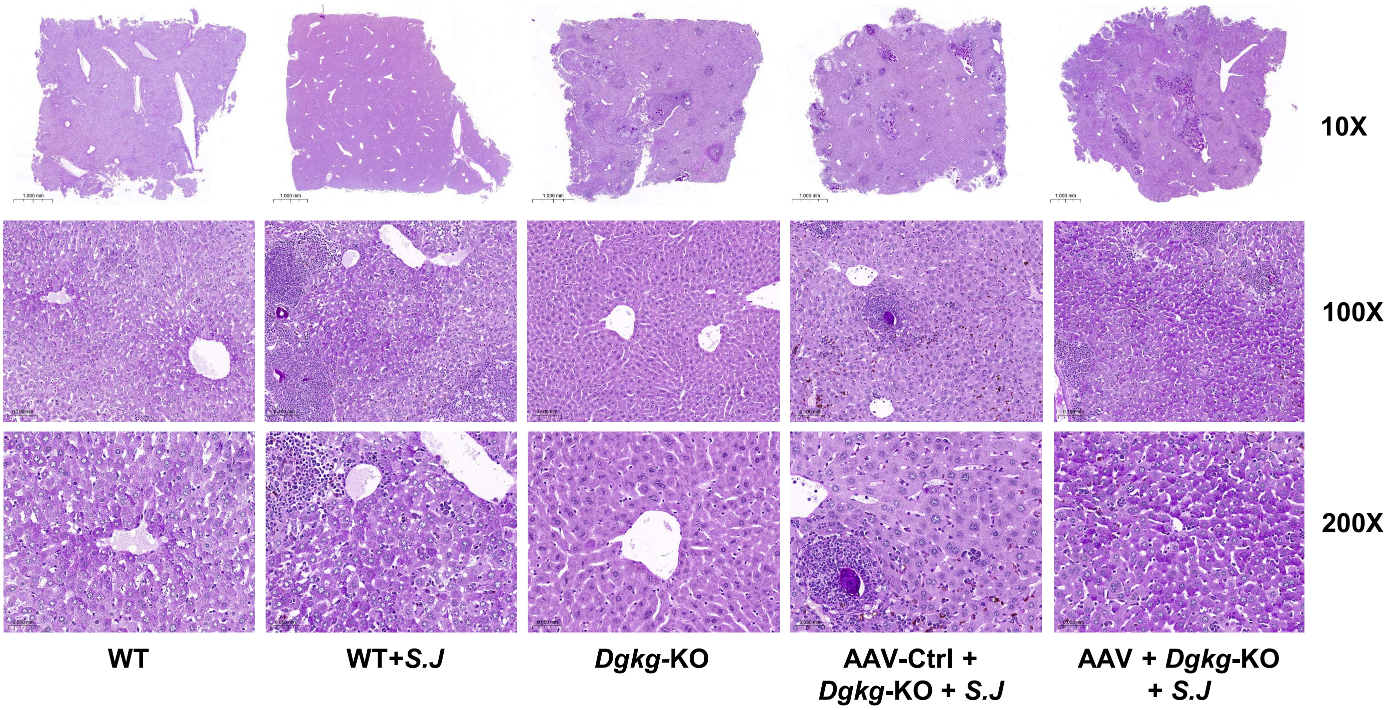


**Supplementary Fig. 9 | Overexpression of DGKG aggravates hepatic glycogen deposition in mice infected with *S. japonicum*.**
